# *Bifidobacterium longum* modifies a nutritional intervention for stunting in Zimbabwean infants

**DOI:** 10.1101/2024.01.18.24301438

**Authors:** Ethan K Gough, Thaddeus J Edens, Lynnea Carr, Ruairi C Robertson, Kuda Mutasa, Robert Ntozini, Bernard Chasekwa, Hyun Min Geum, Iman Baharmand, Sandeep K Gill, Batsirai Mutasa, Mduduzi N N Mbuya, Florence D Majo, Naume Tavengwa, Freddy Francis, Joice Tome, Ceri Evans, Margaret Kosek, Andrew J Prendergast, Amee R Manges, the Sanitation Hygiene Infant Nutrition Efficacy (SHINE) Trial Team

## Abstract

Child stunting is an indicator of chronic undernutrition and reduced human capital. Small-quantity lipid-based nutrient supplements (SQ-LNS) has been widely tested to reduce stunting, but has modest effects. The infant intestinal microbiome may contribute to stunting, and is partly shaped by mother and infant histo-blood group antigens (HBGA). We investigated whether mother-infant fucosyltransferase status, which governs HBGA, and the infant gut microbiome modified the impact of SQ-LNS on stunting at age 18 months among Zimbabwean infants in the SHINE Trial (NCT01824940). We found that mother-infant fucosyltransferase discordance and *Bifidobacterium longum* modified SQ-LNS efficacy. Infant age-related microbiome shifts in *B. longum* subspecies dominance from *infantis*, a proficient human milk oligosaccharide utilizer, to *suis* or *longum*, proficient plant-polysaccharide utilizers, were partly influenced by discordance in mother-infant FUT2+/FUT3-phenotype, suggesting that a “younger” microbiome at initiation of SQ-LNS reduces its benefits on stunting in areas with a high prevalence of linear growth restriction.

## Introduction

Globally, 21% of children under 5 years of age (149 million) are stunted^1^, defined as having a length or height >2 standard deviations below an age– and sex-matched reference population median^2^. Deficits in linear growth largely accrue from conception to 24 months of age^3,4^, corresponding to the period when normal child growth and development is most rapid. Stunting is associated with reductions in child survival, neurodevelopment, educational attainment, and adult economic productivity^5–7^.

A broadly tested intervention to reduce stunting has been provision of small-quantity lipid-based nutrient supplements (SQ-LNS) to improve infant and young child feeding (IYCF) starting at 6-months (mo) of age, which is the recommended time for introduction of complementary foods. Randomized controlled trials (RCT) of SQ-LNS provision to infants have shown small reductions in stunting (12% relative reduction), but effects have varied^8,9^. Evidence to support other nutrition interventions, which address the underlying determinants of stunting during the first 1000 days of life, starting from conception, is also limited^10^. Elucidating the reasons for the limited impact of nutritional interventions that are designed to alleviate stunting is crucial to the development of more effective strategies.

Recently, the human microbiome has been shown to impact infant health^11^, and studies suggest a role of the intestinal microbiome in child growth, particularly ponderal growth^12,13^. The prevailing view holds that intestinal colonization with bacteria begins at birth, after which the microbiome progresses through a succession of identifiable shifts in composition and functional capacity^11^ that correspond closely with infant age and developmental stage^14^. Deviations from an age-appropriate composition may be associated with poor health outcomes^14–17^, including undernutrition in low resource settings^14,18–20^. However, results vary^21–25^ and evidence for a causal effect of the intestinal microbiome on growth comes predominantly from animal models. In these experiments, a combination of commensal bacteria and a nutrient-poor diet produce a synergistic effect on growth faltering. The detrimental effect on growth was worse when certain commensal bacteria, which can act as pathobionts (e.g. *Bacteroides thetaiotaomicron* and *Bacteroides fragilis*), were present in the gut along with pathogenic species^26,27^.

The gut microbiome in infants is influenced by breastfeeding, with differences persisting beyond age 6mo^16^. Breastfeeding-associated differences are partly driven by differences in human milk oligosaccharide (HMO) composition, which are metabolized by specific commensal bacteria and thereby influence the growth and activity of bacterial populations in the infant gut, in particular, Bifidobacterium^28,29^. In addition, HMOs in combination with commensal gut bacteria have been shown to improve growth in animal models of undernutrition^30^. Active maternal α-1,2-fucosyltransferase (FUT2) and α-1,3-fucosyltransferase (FUT3) genes are key determinants of HMO composition^31–33^. These genes encode enzymes which catalyze addition of fucose to the disaccharides that serve as precursors to host glycan production^34,35^. Individuals with at least one functional FUT2 or FUT3 allele produce fucosylated histo-blood group antigens (HBGA) and are called “secretors” or “Lewis-positive”, respectively;^35^ by contrast, individuals lacking two functional FUT2 or FUT3 alleles don’t produce fucosylated HBGAs, and are termed “non-secretors” or “Lewis-null”, respectively^35^. These secretor and Lewis phenotypes act in concert to synthesize a variety of HBGAs (Figure S1)^34,35^. In addition to the impact of maternal FUT2 or FUT3 status on the infant gut microbiome via their influence on HMO composition, host FUT2 and FUT3 phenotypes also determine tissue surface HBGA expression in the gut, which may also affect the microbiome through the availability host glycans and competition for adhesion sites^36–40^.

There is growing interest in the moderating effect of the gut microbiome on nutritional interventions. Recent RCTs showed that the impact of dietary interventions for weight reduction on metabolic health outcomes is modified up to 4-fold by microbiota composition^41–44^. Effect modification of host diet by the gut microbiota has also been investigated in observational studies, which report that microbiome composition modified the association between diet and biomarkers of metabolic syndrome by up to 2-fold^45–47^. Gut microbiota composition, therefore, may also be an important modifier of dietary interventions on infant health. Only one study to date investigated the gut microbiota as an effect modifier of SQ-LNS on infant stunting, and reported limited evidence of effect-modification^48^. However, several studies have reported a synergistic effect of microbiome composition and diet on undernutrition phenotypes using animal models^26,27,49,50^.

Here we aimed to determine the effect of infant gut microbiome composition and functional capacity on efficacy of an IYCF intervention that included SQ-LNS to reduce stunting (primary outcome) and improve length-for-age z-score (LAZ) (secondary outcome) at age 18mo. We hypothesized that the efficacy of IYCF, when started at age 6mo, on infant linear growth is modified by mother and infant FUT2 and FUT3 phenotype, as an drivers of variation in gut microbiome composition, and by the infant gut microbiome. We tested this hypothesis using data from HIV-unexposed infants enrolled in the Sanitation Hygiene Infant Nutrition Efficacy (SHINE) trial conducted in rural Zimbabwe, in which the IYCF intervention modestly increased LAZ by 0.16 standard deviations and reduced stunting by 20% at age 18mo^51^. We found that mother-infant FUT2 and FUT3 phenotype, where the mother was FUT2+/FUT3-but the infant was not, was associated with greater IYCF reduction of stunting at 18mo. Greater impact of IYCF on stunting was also associated with infant microbiome species maturation, characterized by a shift away from *Bifidobacterium longum* carriage. *B. longum*-dominant microbiome composition was characterized by *B. longum* strains that were most similar to subspecies *infantis*, a proficient HMO utilizer. In addition, *B. longum* abundance and odds of detecting different *B. longum* strains were explained by infant age and differences between mother-infant pairs in FUT2+/FUT3-phenotype.

## Results

### Mother-infant FUT2+/FUT3-phenotype discordance modifies the effect of IYCF on stunting at 18mo

In a substudy of the SHINE trial, we assessed FUT2 and FUT3 status in mothers and infants from saliva samples. To determine the impact of maternal and infant HBGA on IYCF efficacy, we investigated whether concordance in secretor (FUT2) and Lewis (FUT3) phenotypes between mother and infant pairs modified the effect of IYCF on stunting or LAZ at 18mo. We defined these paired mother-infant phenotypes using combinations of FUT2 and FUT3 status (as presented in Table S1) to more precisely reflect the potential for joint activity of these enzymes in HBGA production (as presented in Figure S1). Each of these paired mother-infant FUT2/FUT3 phenotypes were classified as *both* (if mother and infant shared the same phenotype), *none* (if neither had the phenotype), *infant only* (if the infant had the phenotype but the mother did not) or *mother only* (if the infant did not have the phenotype but the mother did). We fitted multivariable regression models that included terms for the interaction of IYCF and each paired FUT2/FUT3 phenotype between mother and infant, as well as prespecified covariates from the 6mo follow-up visit when IYCF was started, with stunting status at the 18mo visit as the dependent variable. We fitted separate models for each combination of paired mother-infant FUT2/FUT3 phenotype presented in Table S1 and Figure S1. These models were restricted to 792 infants who had both maternal and infant FUT2 and FUT3 status ascertained (Figure S2). Analyses were repeated with LAZ at the 18mo visit as the dependent variable.

Amongst infants who were randomized to IYCF, those who were in the *mother only* FUT2+/FUT3-group (13.5%) had a probability of stunting at 18mo that was –33.0% lower (95%CI:-55.0%,-10.0%) than infants in the *both* FUT2+/FUT3-group (11.1%) (Table 1). The *infant only* (11.1%) and *none* (64.3%) FUT2+/FUT3-groups, and other mother-infant FUT2 and FUT3 phenotype combinations (Table S1, Figure S1), did not show evidence of effect modification of IYCF on stunting (Table S2), and there was no evidence of effect modification on LAZ at 18mo after FDR-adjustment for multiple testing (Table S3). Overall, our results indicate that discordance between mothers and infants in the FUT2+/FUT3-phenotype, whereby mothers had the phenotype and infants didn’t have the phenotype, was associated with increased efficacy of IYCF to reduce stunting but not to increase LAZ.

**Table 1.**
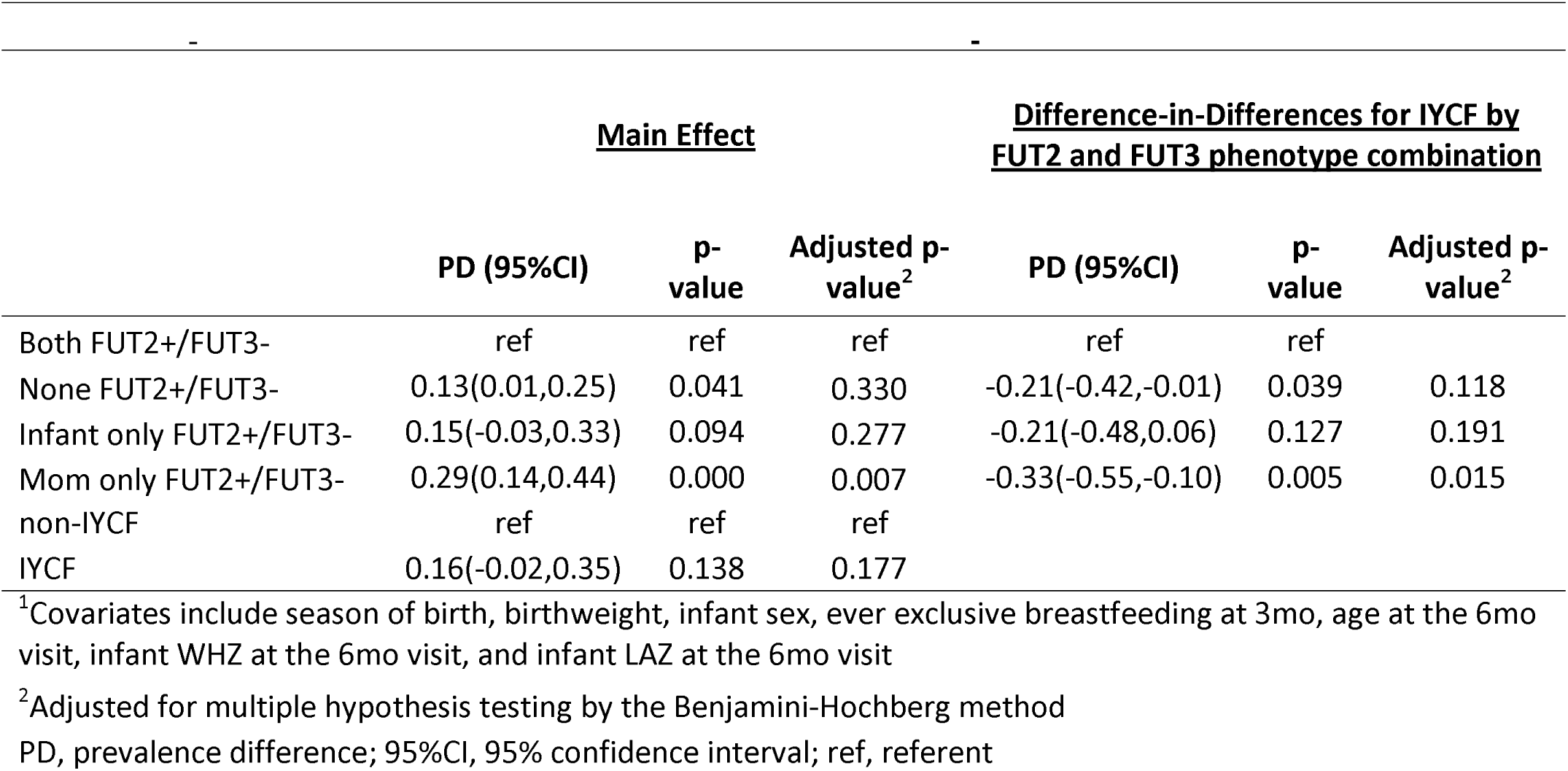
Multivariable regression model^1^ to estimate modification of IYCF on stunting at 18mo by mother-infant Lewis-null secretor phenotype discordance among 792 infants in whom mother and infant FUT2 and FUT3 status was ascertained.

|  | <u>Main Effect</u> |  |  | <u>Difference-in-Differences for IYCF by FUT2 and FUT3 phenotype combination</u> |  |  |
| --- | --- | --- | --- | --- | --- | --- |
|  | PD (95%CI) | p-value | Adjusted p-value<sup>2</sup> | PD (95%CI) | p-value | Adjusted p-value<sup>2</sup> |
| Both FUT2+/FUT3- | ref | ref | ref | ref | ref |  |
| None FUT2+/FUT3- | 0.13(0.01,0.25) | 0.041 | 0.330 | -0.21(-0.42,-0.01) | 0.039 | 0.118 |
| Infant only FUT2+/FUT3- | 0.15(-0.03,0.33) | 0.094 | 0.277 | -0.21(-0.48,0.06) | 0.127 | 0.191 |
| Mom only FUT2+/FUT3- | 0.29(0.14,0.44) | 0.000 | 0.007 | -0.33(-0.55,-0.10) | 0.005 | 0.015 |
| non-IYCF | ref | ref | ref |  |  |  |
| IYCF | 0.16(-0.02,0.35) | 0.138 | 0.177 |  |  |  |

<sup>1</sup>Covariates include season of birth, birthweight, infant sex, ever exclusive breastfeeding at 3mo, age at the 6mo visit, infant WHZ at the 6mo visit, and infant LAZ at the 6mo visit
<sup>2</sup>Adjusted for multiple hypothesis testing by the Benjamini-Hochberg method
PD, prevalence difference; 95%CI, 95% confidence interval; ref, referent

### Infant gut microbiome composition modifies the effect of the IYCF intervention on stunting at 18mo

Given our finding that infants whose mothers were FUT2+/FUT3-, while the infant was not, had greater reductions in stunting by IYCF; and considering the potential importance of FUT2/FUT3 phenotypes on early infant microbiome composition via variation in maternal HMO composition and infant gut epithelial HBGA expression, we investigated whether infant gut microbiota species maturation showed evidence for modification of IYCF efficacy on stunting or LAZ at 18mo. First, we used 354 infant metagenomes from 172 infants collected from 1-18mo of age to fit a constrained Principal Coordinates Analysis (PCoA) model that included infant age at stool collection and three dummy variables for mother-infant FUT2+/FUT3-status representing the *none*, *infant only*, and *mother only* groups with the *both* group as the referent (see next section for details). Thus, we derived four PCoA axis scores from this model representing variation in microbiome composition due to infant age and explained by *none*, *infant only*, or *mother only* FUT2+/FUT3-status compared to *both*. Next, we fitted a multivariable regression model that included an IYCF-by-PCoA axis 1 score interaction term and prespecified covariates. The regression model was restricted to 53 infants who had a fecal specimen collected at the 6mo follow-up visit when IYCF started and used their covariate values at the 6mo visit (Figure S2). Stunting status at 18mo was used as the dependent variable. We repeated our analyses using PCoA axis 2 to 4 scores. Finally, we fitted regression models replacing stunting status with LAZ at 18mo as the dependent variable.

Greater PCoA axis 1 scores represented species turnover with increasing infant age (Figure S3), thus reflecting microbiota maturation, while greater PCoA axis 2 scores represented changes in species composition associated with mother-infant FUT2+/FUT3-phenotypes (Figure S3). PCoA axis 1 and 2 scores showed evidence of interaction with IYCF on stunting at 18mo (Table 2), but not LAZ (Table S4). Amongst infants randomized to IYCF, those with higher PCoA axis 1 scores at age 6mo were less likely to be stunted at 18mo compared to infants with lower PCoA axis 1 scores (difference-in-differences –76.0% [95%CI: –99.0%,-32.0%, adjusted p=0.003) (Table 2). In contrast, infants randomized to IYCF with higher PCoA axis 2 scores at age 6mo were more likely to be stunted at 18mo compared to infants who had lower PCoA axis 2 scores (difference-in-differences, 14.0% [95%CI: 7.0%,21.0%], adjusted p=0.001) (Table 2). Taken together, our findings showed that infant age-related microbiome species maturation was associated with greater reduction in the probability of stunting at 18mo by IYCF, indicating greater efficacy of the intervention; while microbiome species composition related to mother-infant FUT2+/FUT3-phenotype was associated with lesser reduction in stunting, suggesting variation in microbiome composition due to these infant characteristics is an important determinant of SQ-LNS efficacy to reduce stunting.

**Table 2.**
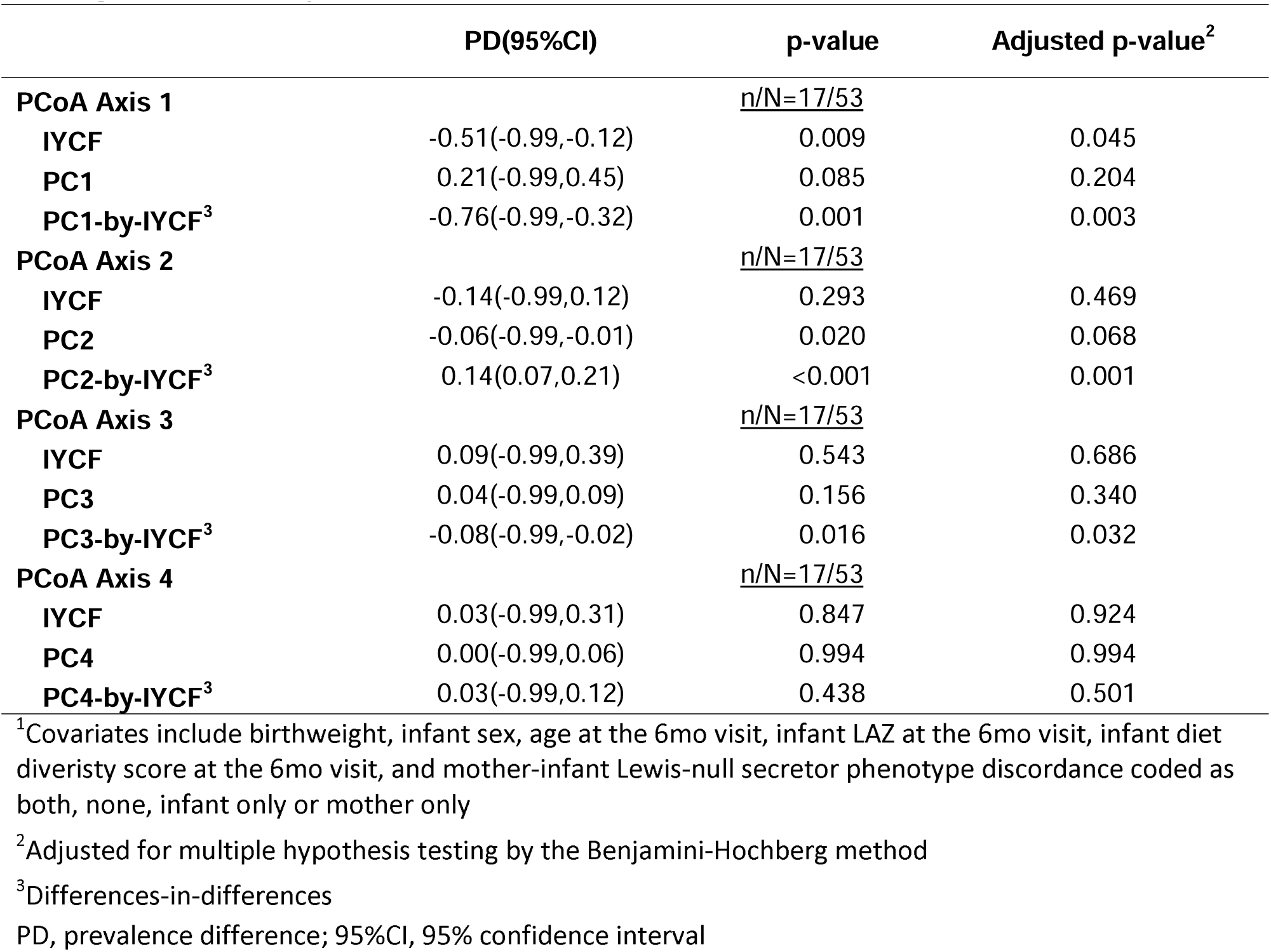
Multivariable regression models^1^ to estimate modification of IYCF on stunting at 18mo by infant gut microbiome species turnover in 53 infants.

### Infant age and mother-infant FUT2+/FUT3-phenotype explain shifts in microbiome species composition, in particular, Bifidobacterium longum

To investigate sources of variation in infant intestinal microbiome composition and derive interpretable measures of species turnover, we performed constrained PCoA of Bray-Curtis dissimilarities with permutational analysis of variance using distance matrices for hypothesis testing. Infant age at stool collection explained 12.6% (ADONIS2 p=0.001) of the variability in microbiome composition (Table S5). Notably, exclusive breastfeeding (EBF) at 3mo was not significantly associated (*R*^2^=0.346, ADONIS2 p=0.483) with microbiome composition (Table S5). However, 93.2% of mothers in these analyses reported exclusive breastfeeding (Table S6). Thus, there may not have been sufficient variability in infant breastfeeding practices in this cohort to identify EBF-associated differences between infant microbiomes. Also, inclusion of specimens collected throughout the follow-up period in our PCoA models may have obscured associations with EBF, since EBF is only recommended up to age 6mo.

We then fitted a fully adjusted multivariable constrained PCoA model that included both infant age at stool collection and mother-infant FUT2+/FUT3-phenotype. We retained the latter variable to capture variation in microbiota composition associated with mother-infant FUT2+/FUT3-status after controlling for infant age, because of our *a priori* aim to investigate whether microbiome variation due to mother and infant FUT2/FUT3 phenotype was a modifier of the IYCF intervention, and our finding that the *mother only* FUT2+/FUT3-group was associated with a modified intervention effect. In the full model, age explained 12.8% (ADONIS p=0.001) and mother-infant FUT2+/FUT3-phenotype explained 0.81% (ADONIS p=0.323) of the variation in microbiome composition (Table S5), indicating that infant age was the main driver of microbiome-wide changes in species composition in this cohort.

Age-related maturation in species composition were characterized by decreased relative abundance of *B. longum*, and increased relative abundance of *Prevotella copri*, *Faecalibacterium prausnitzii*, *Dorea longicatena*, and *Dorea formicigenerans* (Figure S4). Mother-infant FUT2+/FUT3-discordance was characterized by increased relative abundance of *B. longum* in the *none* and *infant only* groups (Figure S5-S7).

In combination with our IYCF-by-PCoA effect modification models (see previous section), these findings indicate that infant microbiota species maturation characterized by a shift away from a microbiome dominated by *B. longum* (i.e. greater PCoA axis 1 score) was associated with a reduction in the probability of stunting at age 18mo in infants randomized to IYCF (Table 2). In contrast, species composition associated with mother-infant FUT2+/FUT3-phenotype (i.e. greater PCoA axis 2 score), which was characterized by higher *B. longum* abundance, showed evidence of reduced impact of IYCF on stunting (Table 2).

### Bifidobacterium longum relative abundance modifies the effect of the IYCF intervention on stunting at 18mo

To determine if differences in the relative abundances of specific microbiome species that characterized the constrained PCoA axes were also associated with significant modification of IYCF on stunting or LAZ at 18mo, we fitted multivariable regression models that included IYCF-by-species relative abundance interaction terms and prespecified covariates as described in the previous sections. These models were restricted to 53 infants who had a fecal specimen collected at the 6mo follow-up visit when IYCF started and used their covariate values at that visit (Figure S2). We fitted a separate model for each microbiome species of interest, defined as those strongly associated with PCoA axis 1 to 4 scores (i.e. loadings > 0.5 or < –0.5). Infants randomized to IYCF who had greater relative abundance of *B. longum* when the intervention started were more likely to be stunted at age 18mo compared to infants with lower relative abundance of *B. longum* (differences-in-differences 50.0% [95%CI: 26.0%,74.0%], adjusted p<0.001) (Table 3). No other species-level interactions with IYCF on stunting were identified (Table 3). Infants randomized to IYCF with greater *B. longum* relative abundance also had smaller LAZ at 18mo compared to infants with lower *B. longum* abundance; however, this association was not statistically significant after FDR-adjustment (differences-in-differences –0.88[95%CI: –1.45,-0.31], adjusted p=0.074) (Table S7). Overall, our findings indicate that greater relative abundance of *B. longum* in the infant gut microbiome at initiation of the IYCF intervention was associated with less efficacy of the IYCF intervention to reduce stunting at age 18mo.

**Table 3.** Multivariable regression models^1^ to estimate modification of IYCF on stunting at 18mo by infant gut microbiome species in 53 infants.

|  | PD(95%CI) | p-value | Adjusted p-value<sup>2</sup> |
| --- | --- | --- | --- |
| Bifidobacterium longum |  | <u>n/N=17/53</u> |  |
| IYCF | -0.16(-0.43,0.10) | 0.232 | 0.976 |
| B.longum | -0.24(-0.43,-0.05) | 0.014 | 0.068 |
| B.longum-by-IYCF<sup>3</sup> | 0.50(0.26,0.74) | <0.001 | <0.001 |
| Bifidobacterium pseudocatenulatum |  | <u>n/N=17/53</u> |  |
| IYCF | -0.54(-1.02,-0.06) | 0.029 | 0.304 |
| B.pseudocatenulatum | 1.82(0.48,3.15) | 0.008 | 0.051 |
| B.pseudocatenulatum-by-IYCF<sup>3</sup> | -2.00(-3.33,-0.68) | 0.003 | 0.014 |
| Escherichia coli |  | <u>n/N=17/53</u> |  |
| IYCF | 0.04(-0.25,0.33) | 0.774 | 1.000 |
| E.coli | 0.01(-0.10,0.13) | 0.823 | 0.859 |
| E.coli-by-IYCF<sup>3</sup> | -0.07(-0.21,0.08) | 0.362 | 0.592 |
| Dorea longicatena |  | <u>n/N=17/53</u> |  |
| IYCF | 0.02(-0.27,0.31) | 0.896 | 1.000 |
| D.longicatena | -0.02(-0.13,0.09) | 0.725 | 0.859 |
| D.longicatena-by-IYCF<sup>3</sup> | 0.03(-0.39,0.44) | 0.898 | 0.898 |
| Dorea formicigenerans |  | <u>n/N=17/53</u> |  |
| IYCF | -0.18(-0.37,0.01) | 0.062 | 0.324 |
| D.formicigenerans | -0.18(-0.33,-0.03) | 0.018 | 0.073 |
| D.formicigenerans-by-IYCF<sup>3</sup> | -0.50(-1.00,-0.00) | 0.048 | 0.145 |

<sup>1</sup>Covariates include birthweight, infant sex, age at the 6mo visit, infant LAZ at the 6mo visit, infant diet diversity score at the 6mo visit, and mother-infant Lewis-null secretor phenotype discordance coded as both, none, infant only or mother only
<sup>2</sup>Adjusted for multiple hypothesis testing by the Benjamini-Hochberg method
<sup>3</sup>Differences-in-differences
PD, prevalence difference; 95%CI, 95% confidence interval

### Infants randomized to IYCF in the mother only FUT2+/FUT3-group or with lower B. longum relative abundance were spared from more severe growth faltering

To investigate whether these reductions in IYCF efficacy were due to differences in the degree or timing of growth faltering, we plotted (i) LAZ growth trajectories and (ii) LAZ velocities (sd/mo) from 6-18mo of age, by both IYCF arm and paired mother-infant FUT2+/FUT3-phenotype. Infants randomized to IYCF in the *mother only* group had a less steep decline in LAZ trajectories on average compared to infants in the *mother only* group randomized to no IYCF (Figure 1). In addition, although all groups of infants had negative 6-18mo LAZ velocities on average, infants in the *mother only* group who were randomized to IYCF had higher LAZ velocity (−0.02 sd/mo,95%CI[-0.05,0.00]) compared to those in the no IYCF group (−0.08 sd/mo, 95%CI[-0.11,-0.05]) (Figure 1). Importantly, infants in the *mother only* group also benefitted more from the IYCF intervention in our interaction models (Table 1).

We repeated these exploratory analyses to compare LAZ trajectories and LAZ velocities by both IYCF arm and *B. longum* at the 6mo visit stratified above or below the relative abundance median. Infants randomized to IYCF who were above the median relative abundance of *B. longum* had declining growth trajectories on average, while infants below the median did not (Figure 2). Furthermore, the mean 6-18mo LAZ velocity was negative in both *B. longum* groups, but trended toward being higher in the group with relative abundance below (−0.02 sd/mo,95%CI[-0.07,0.03]) compared to above (−0.08 sd/mo 95%CI[-0.12,-0.03]) the median; however, the 95% confidence intervals overlapped. In contrast, among infants randomized to no IYCF, both those with a relative abundance above and those with a relative abundance below the median at the 6mo visit had declining growth trajectories (Figure 2) with similar average negative LAZ velocities (Figure 2).

**Figure 1.**
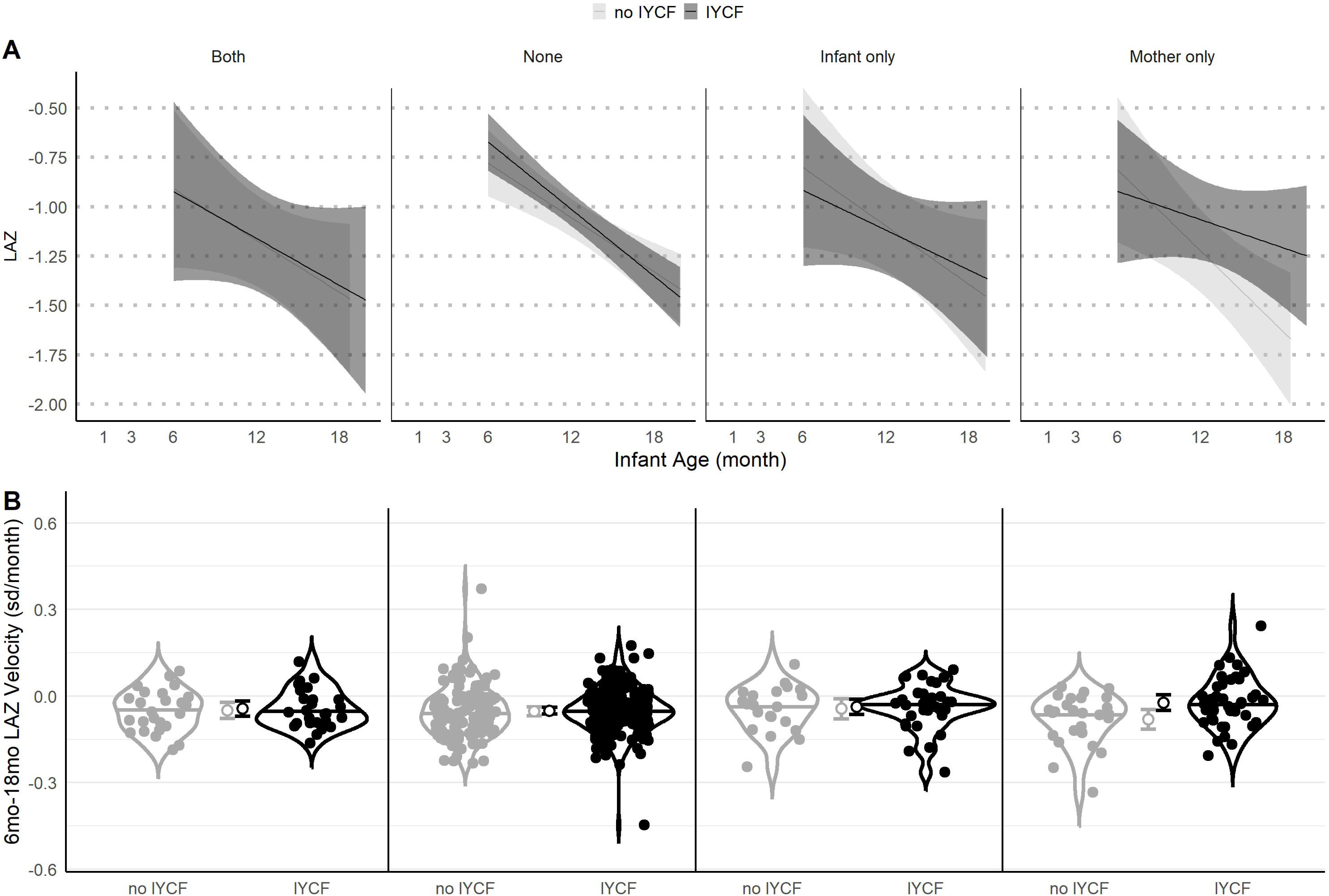
Infant growth trajectories and growth velocities vary by IYCF arm and mother-infant FUT2+/FUT3-phenotype. (A) LAZ by infant age from 1-18mo (n=792), stratified by IYCF arm (light grey, no IYCF; dark grey, IYCF) and mother-infant FUT2+/FUT3-phenotype (*both*, *none*, *infant only* or *mother only*). Lines illustrate average trajectories. Shaded areas are 95% confidence bands. Infants randomized to IYCF in the *mother only* group had a less steep decline in LAZ trajectories on average compared to infants randomized to no IYCF. (B) Violin plots of LAZ velocity from 6-18mo of age, stratified by IYCF arm (light grey, no IYCF; dark grey, IYCF) and mother-infant FUT2+/FUT3-phenotype (*both*, *none*, *infant only* or *mother only*). Open circles with error bars in the center of each panel indicate mean LAZ velocity and 95%CIs. All groups of infants had negative LAZ velocities on average, but infants in the *mother only* group who were randomized to IYCF had higher LAZ velocity on average, contributing to a less steep decline in LAZ over time and a smaller proportion of infants stunted at 18mo.

**Figure 2.**
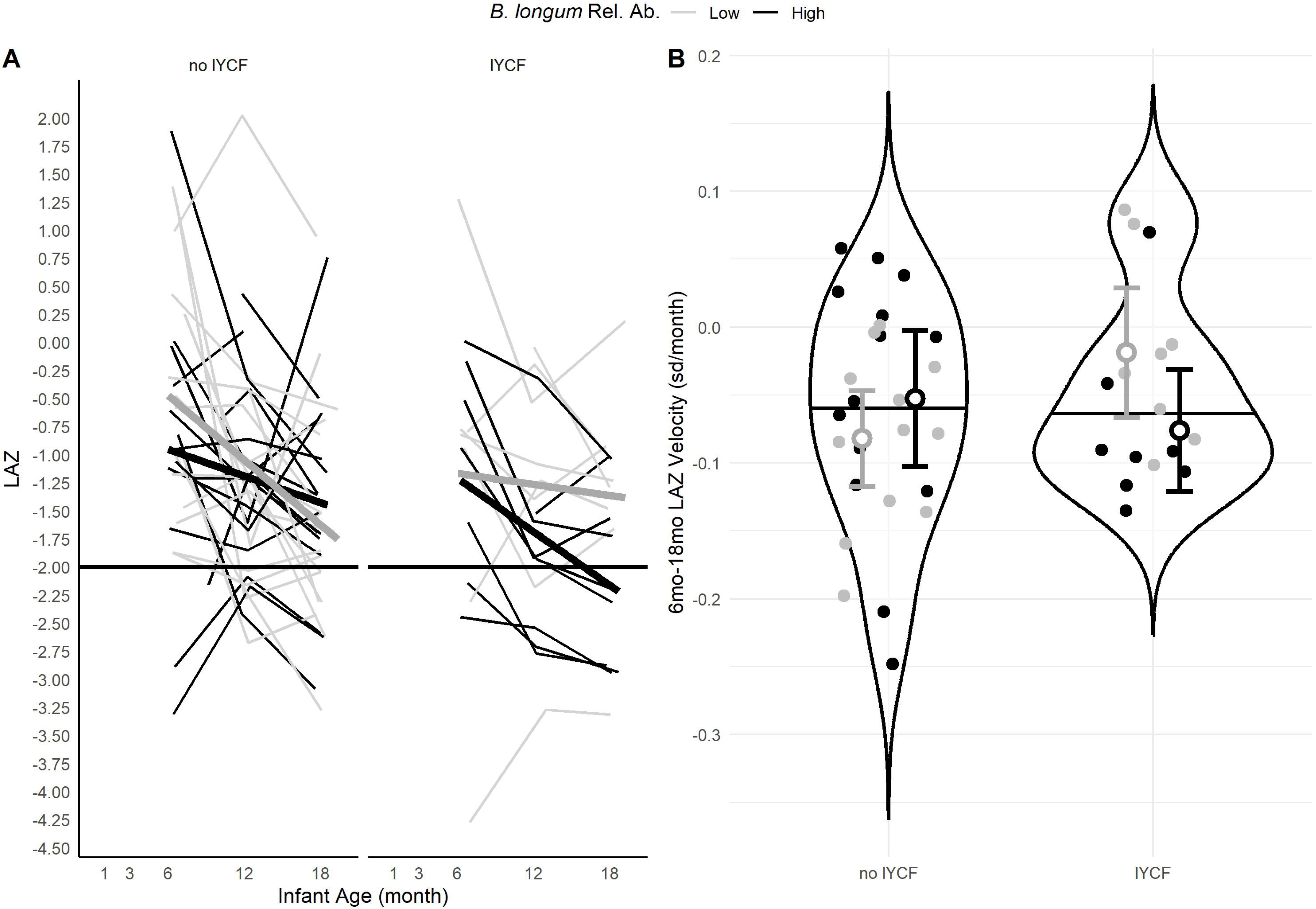
Infant growth trajectories and growth velocities vary by IYCF arm and infant gut *Bifidobacterium longum* relative abundance. (A) LAZ by infant age from 1-18mo (n=53), stratified by IYCF arm and *Bifidobacterium longum* relative abundance (light grey, ≤median relative abundance; dark grey, >median relative abundance). Thin lines illustrate individual trajectories. Thick lines show average trajectories. Infants randomized to IYCF who were above the median relative abundance had a declining growth trajectory on average, while infants below the median did not. (B) Violin plots of LAZ velocity from 6-18mo of age, stratified by IYCF arm (light grey, no IYCF; dark grey, IYCF) and mother-infant FUT2+/FUT3-phenotype (*both*, *none*, *infant only* or *mother only*). Open circles with error bars indicate mean LAZ velocity and 95%CIs. All groups of infants had negative LAZ velocities on average. However, LAZ velocity trended toward being higher in the group with relative abundance below the median. The 95% confidence intervals overlap, but the higher LAZ velocity contributed to a less steep decline in LAZ over time and a smaller proportion of stunted infants at 18mo.

Overall, these results indicate that the differences in IYCF efficacy by mother-infant FUT2+/FUT3-phenotype and *B. longum* relative abundance can be explained by differences in LAZ velocity. Infants in the *mother only* FUT2+/FUT3-group and infants with lower *B. longum* relative abundance at 6mo were spared from more severe LAZ declines, which contributed to a lower probability of stunting at 18mo, even when the differences in velocity were not statistically significant.

### Bifidobacterium longum strains most similar to subspecies infantis dominate the early infant gut microbiome

Next, we aimed to identify whether infants carried different strains of *B. longum* using a pangenome approach. We used PanPhlan3.0^52^ to generate UniProt gene family profiles of dominant *B. longum* strains from 218 infant gut metagenomes, generated from 136 infant, with sufficient coverage of *B. longum* (Figure S2). To assess similarities between SHINE *B. longum* strains and previously characterized subspecies, we also included UniProt gene family profiles produced by PanPhlan for 118 reference strains. We performed PCoA using Jaccard dissimilarities calculated from these gene family presence or absence profiles. Three clusters of *B. longum* strains were identifiable by visualization of ordination plots (Figure 3 and S8). We, therefore, performed hierarchical clustering of these Jaccard dissimilarities to delineate three distinct strain clusters and place each strain into the most appropriate cluster (Figure S8). Subspecies *infantis* reference strains predominantly grouped with the largest cluster (hereafter called the *infantis* cluster) and included 15 *infantis* reference strains and 255 SHINE strains (Figure 3). Subspecies *longum* reference strains predominantly grouped with the second largest cluster (*longum* cluster), including 29 *longum*, 3 *infantis*, 64 *unclassified* subspecies strains and 14 SHINE strains. The remaining cluster included two subspecies *suis*, one subspecies *longum*, 4 *unclassified* subspecies reference strains and 15 SHINE strains (*suis* cluster) (Figure 3 and Figure S8).

**Figure 3.**
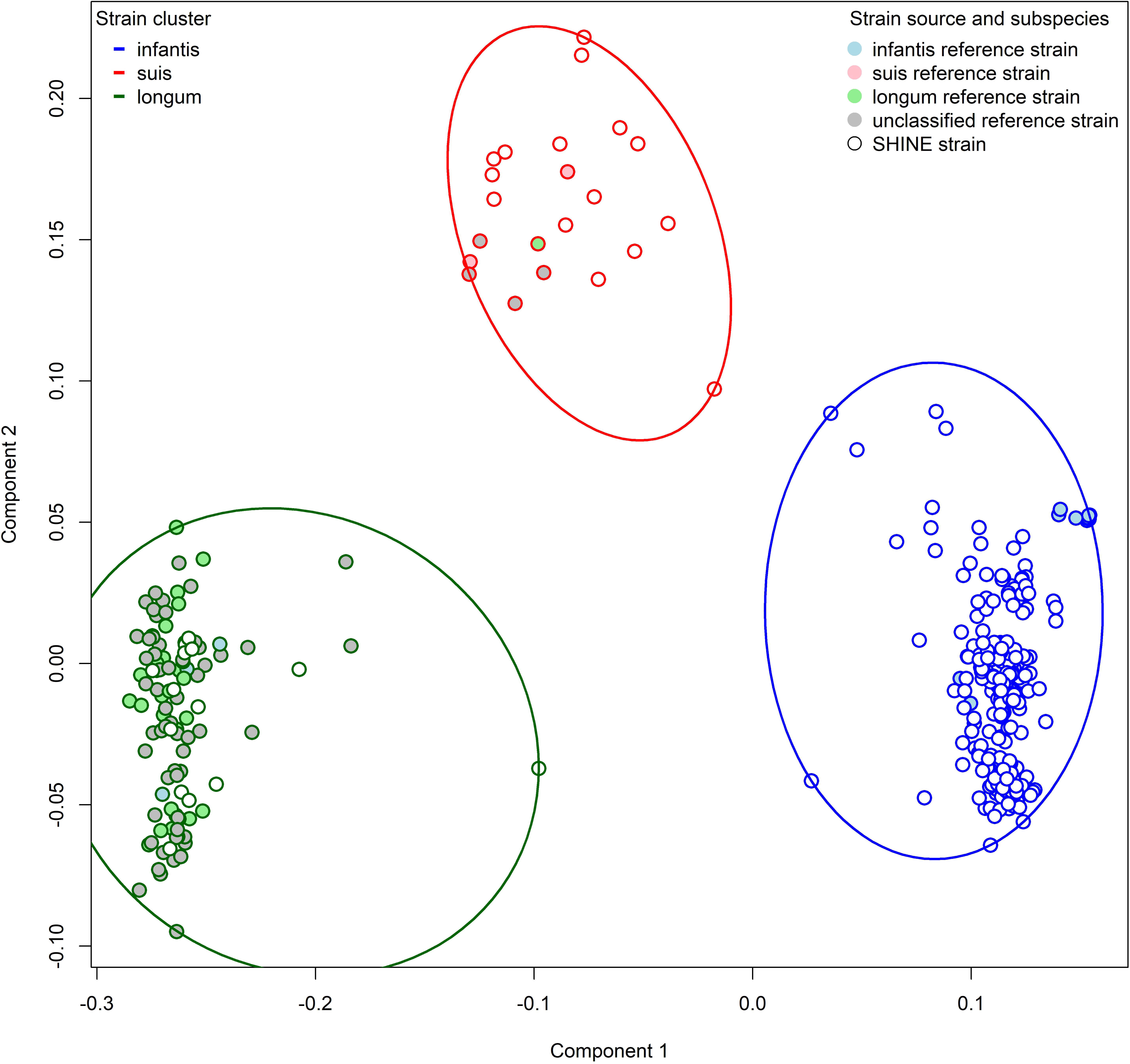
*Bifidobacterium longum* strains most similar to subspecies *infantis* dominate the early infant gut microbiome. Ordination plots of PCoA with Jaccard dissimilarities calculated using the pangenome profiles of dominant *Bifidobacterium longum* strains in fecal specimens (N=284) generated using PanPhlan3.0. Three clusters of *B. longum* strains were identifiable. Individual strains and indicated by small circles. Those in the same cluster are enclosed by a large ellipse and are differentiated by both the color of the ellipse and the border color of the small circle (blue, *infantis*; dark green, *longum*; red, *suis*). Filled small circles indicate reference strains (light blue, *infantis*; pink, *suis*, light green, *longum*; grey, *unclassified*). Open small circles indicate SHINE strains. Subspecies *infantis* reference strains predominantly grouped with the largest cluster (blue ellipse, *infantis* cluster). Subspecies *longum* reference strains predominantly grouped with the second largest cluster (dark green ellipse, *longum* cluster). The remaining cluster (red ellipse, *suis* cluster) grouped with subspecies *suis* and *longum*.

### Infant gut microbiome maturation undergoes a transition in which Bifidobacterium longum strains most similar to subspecies infantis become less prevalent

To investigate predictors of *B. longum* relative abundance in infants over time, we fitted a longitudinal multivariable zero-inflated mixed-effects model, using 225 specimens from 87 infants with prespecified covariate data. Covariates included infant age at specimen collection, sex, EBF at 3mo, minimum infant dietary diversity at specimen collection, and mother-infant FUT2+/FUT3-phenotypes (Figure S2). Infant age and mother-infant FUT2+/FUT3-phenotype were significant predictors of *B. longum* relative abundance. Each one month increase in infant age was associated with a 0.91-fold decrease (95%CI:0.90,0.92, p<0.001) in *B. longum*, and the *mother only* FUT2+/FUT3-group had the lowest relative abundance of *B. longum* throughout follow-up (Figure S11) with a 0.71-fold decrease in relative abundance (95%CI:0.59,0.87, p=0.001) relative to the *both* group (Table S9).

Also, to determine predictors of *B. longum* strain cluster detection in infants over time, we fitted multivariable longitudinal logistic regression models using 133 specimens from 70 infants with PanPhlan3.0 output and prespecified covariate data (Figure S2). We fitted a separate model for each strain to estimate its probability of detection. Results were consistent with predictors of *B. longum* relative abundance. Each one month increase in infant age was associated with a 0.85-fold decreased odds (95%CI:0.75,0.96, p=0.008) of the *infantis* cluster (Table S10). At the same time, the *suis* and *longum* clusters increased with age (Table S10 & Figure S12). Female infants had 8.35-fold increased odds of the *infantis* cluster (95%CI:3.17,21.97, p<0.001); and the *infant only* FUT2+/FUT3-group, which had the highest probability of the *infantis* cluster throughout follow-up (Figure S13A), had a 4.66-fold increased odds (95%CI:1.05,20.71, p=0.043) of the *infantis* cluster (Table S10). In contrast, the *mother only* FUT2+/FUT3-group had low probability of carrying an *infantis* cluster strain (Figure S13A & Table S10) and the highest probability of carrying a *suis* cluster strain (Figure S13C).

In summary, *B. longum* decreased with infant age and was lowest among infants in the *mother only* FUT2+/FUT3-group. The *infantis* cluster, which included the dominant *B. longum* strains, also decreased with infant age. Furthermore, *infantis* cluster strains were most likely to be detected in the *infant only* group, and were less likely to be detected in the *mother only* group, among whom *suis* cluster strains were more likely to be detected. Infant age and mother-infant FUT2+/FUT3-phenotype were important determinants of both *B. longum* relative abundance and strain carriage.

### Bifidobacterium longum strains most similar to subspecies infantis were characterized by greater capacity for HMO degradation, uptake, siderophore and antimicrobial biosynthesis

To identify differences in metabolic potential between *B. longum* strain clusters, we used two-sided Fisher’s Exact tests to compare UniProt gene family presence between clusters. We restricted these analyses to the 284 SHINE *Bifidobacterium* pangenomes to make inferences about differences in metabolic potential between SHINE strains. We then performed overrepresentation analyses^53^ of differentially frequent gene families by one-sided Fisher’s Exact test to determine whether they were more likely to be involved in specific GO biological processes^54^, or to function as specific carbohydrate-active enzymes (CAZymes)^55^ or transporters^56^. Gene families that were more common in the *infantis* cluster were more likely to be involved in HMO degradation, and included genes that function as CAZyme Glycoside Hydrolase Family 20 (GH20) (β-N-acetylglucosaminidases, β-N-acetylgalactosamindase, β-6-SO3-N-acetylglucosaminidases, and lacto-N-biosidases), GH29 (fucosidases), GH95 (fucosidases), and GH33 (sialidases) (Figure 4A, Table S8). Conversely, gene families that were more common in the other clusters were more likely to be involved in degradation of plant-derived polysaccharides, including genes that function as GH42 (β-galactosidases), GH51 (L-arabinfuranosidases) and GH127 (β-L-arabinofuranosidase) CAZymes (Figure 4A, Table S8). Similarly, gene families that were more common in the *infantis* cluster were more likely to function as transporters involved in oligosaccharide uptake (TCID 3.A.1.1.59), while those that were less common in the *infantis* cluster were more likely to be involved in uptake of fructose and other sugars (TCID 3.A.1.2.23) (Figure 4A, Table S8). Other differences between strain clusters included greater carriage of gene families among *infantis* cluster strains that are involved in riboflavin biosynthesis, signal transduction, amino acid catabolism and uptake.

**Figure 4.**
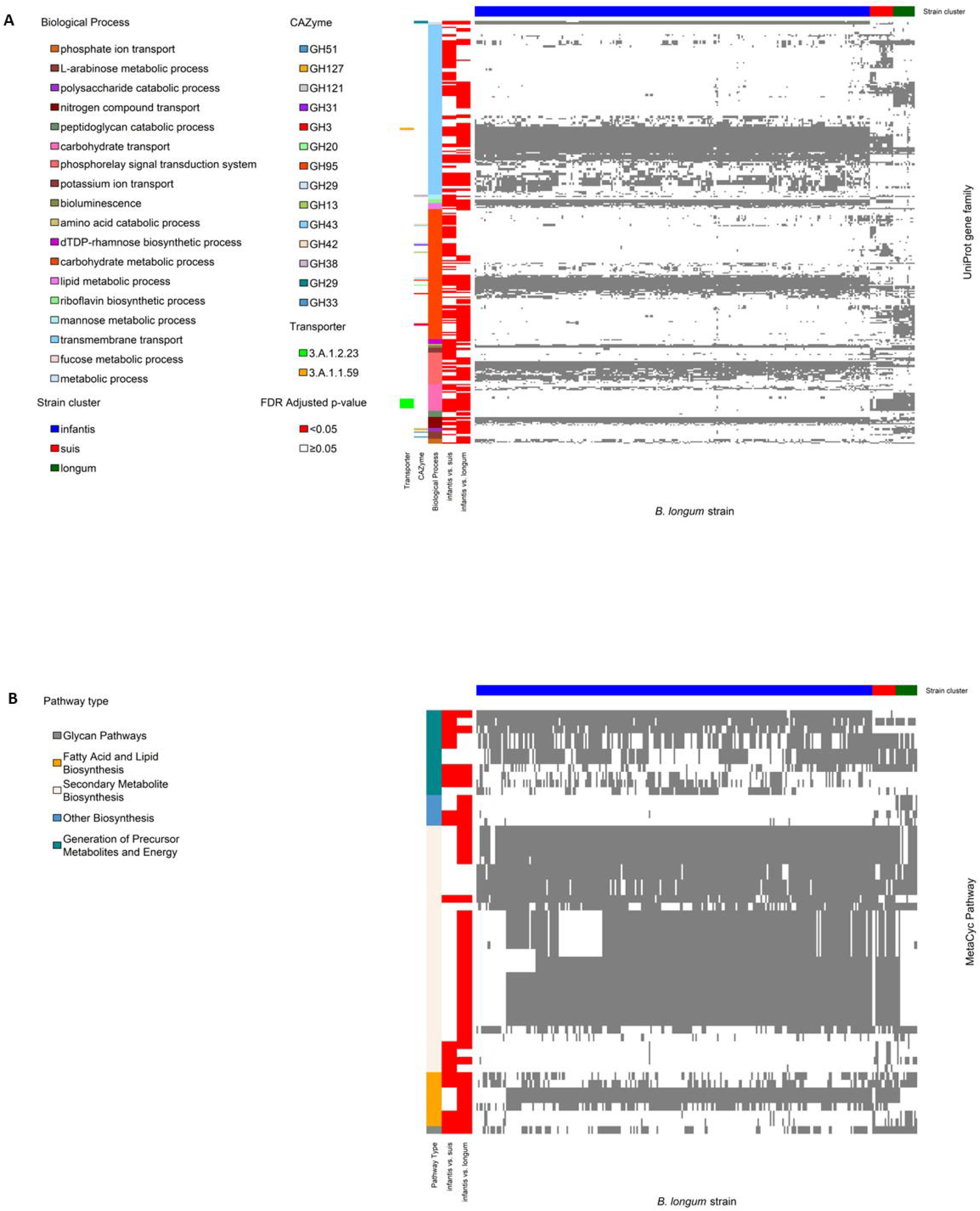
SHINE infant *Bifidobacterium longum infantis* cluster strains are better adapted for human milk oligosaccharide utilization and protection against pathogens than *suis* and *longum* cluster strains. (A) Heatmap of UniProt gene family presence in SHINE infant *Bifidobacterium longum* strains (N=284). UniProt gene family presence is indicated in grey. The horizontal bar at the top indicates strain cluster (blue, *infantis*; dark green, *longum*; red, *suis*). Vertical bars from left to right indicate: UniProt gene families that differed in prevalence between the *infantis* and *longum* cluster by two-sided Fisher’s Exact test after FDR correction (red); UniProt gene families that differed in prevalence between the *infantis* and *suis* cluster by two-sided Fisher’s Exact test after FDR correction (red); Biological Process GO groups; CAZymes; and Transporter class. Only gene families with evidence of overrepresentation in a Biological Process, CAZyme or Transporter class by one-sided Fisher’s Exact Test are presented. (B) Heatmap of MetaCyc pathway presence in SHINE infant *Bifidobacterium longum* strains (N=284). MetaCyc pathway presence is indicated in grey. The horizontal bar at the top indicates strain cluster (blue, *infantis*; dark green, *longum*; red, *suis*). Vertical bars from left to right indicate: MetaCyc pathways that differed in prevalence between the *infantis* and *longum* cluster by two-sided Fisher’s Exact test after FDR correction (red); MetaCyc pathways that differed in prevalence between the *infantis* and *suis* cluster by two-sided Fisher’s Exact test after FDR correction (red); MetaCyc pathway type. Only pathways with evidence of overrepresentation in a pathway type by one-sided Fisher’s Exact Test are presented.

We also identified differences between strain clusters in the frequency of 115 MetaCyc^57^ pathways identified using MinPath^58^ (Figure S10). Pathways that were more common in the *infantis* cluster were more likely to be involved in generation of precursor metabolites and energy, and secondary metabolite biosynthesis (which predominantly included pathways for biosynthesis of siderophores and antimicrobials) (Figure 4B, Table S8). Conversely, the *infantis* cluster was less likely to include pathways involved in glycan metabolism and polymeric compound degradation (e.g. pectin, xylan, and arabinogalactan degradation pathways), and in fatty acid and lipid biosynthesis (Figure 4B, Table S8).

Overall, infant gut microbiomes were dominated by *B. longum* strains that were most similar to subspecies *infantis* in their UniProt gene family profiles. The primary distinction of *B. longum* strains in the *infantis* cluster was greater capacity for HMO degradation and uptake of oligosaccharides than plant-derived polysaccharides. However, strains in the *infantis* cluster also had greater capacity for siderophore and antimicrobial production, and displayed differences in their capacity for riboflavin, fatty acid, and lipid metabolism.

## Discussion

In this study of HIV-unexposed infants enrolled in the SHINE trial in rural Zimbabwe, we tested the hypothesis that mother-infant FUT2/FUT3 phenotype and infant gut microbiome modify the effect of an intervention containing small-quantity lipid based nutrient supplements, on infant stunting and LAZ at 18mo of age. Our goal was to define mechanisms that explain the rather modest effects of SQ-LNS on linear growth, reasoning that maternal-infant HBGA phenotypes might be important given their combined role in shaping the early-life microbiome. We found the following features are associated with greater reduction in stunting at 18mo by IYCF: (i) discordance in mother-infant FUT2+/FUT3-phenotype, where the mother has the phenotype but the infant does not; (ii) changes in microbiome species composition that reflected a shift from a *B. longum*-dominant microbiome to a microbiome with less *B. longum* and a greater abundance of species characteristic of older infants; and (iii) decreased *B. longum* abundance that was associated with paired mother-infant FUT+/FUT3-status. These findings suggest that a persistently “younger” microbiome, characterized by a high abundance of *B. longum* and carriage of *B. longum* strains best suited to HMO metabolism, at the point when complementary foods are introduced into the infant diet, may modify the effects of an intervention that includes SQ-LNS on infant stunting.

The *mother only* FUT2+/FUT3-group showed evidence of a greater reduction in stunting following receipt of one year of the IYCF intervention. Mother-infant FUT2+/FUT3-phenotype was also a significant predictor of *B. longum*, whereby relative abundance was lower throughout follow-up in the *mother only* group. Active maternal FUT2/FUT3 genes are key determinants of milk oligosaccharide composition^31–33^. Human milk specimens form distinct groups based on maternal FUT2^59^ and FUT3 phenotype^31,33^ by principal component analyses of oligosaccharide composition. In particular, FUT2+/FUT3-mothers are a subset of FUT2+ women who form an HMO cluster that is distinct from mothers with other phenotypes, including FUT2+ mothers who are also FUT3+^31,33^. Differences in HMO composition influence growth and activity of Bifidobacterium populations in the infant gut^28,59^. Host FUT2 status may also affect infant gut microbiome composition^36–40^. Gut Bifidobacterium^39^, Bacteroides^36,38^, Faecalibacterium^40^ and Roseburia^40^ are differentially abundant in FUT2+ compared to FUT2-individuals. However, reported differences in microbiome composition by FUT2/FUT3 status have been inconsistent, potentially due to variability in age, dietary patterns, host health, sampled gut sections and methodology between studies^60,614^, and studies predominantly included adults. However, CAZymes found in *B. longum* species that function in HMO degradation (GH29 and GH95) have also been found to function in host intestinal glycan degradation in infants^62^. The evidence, therefore, suggests that the FUT2/FUT3 phenotypes of mother and infant, together, can elicit a strong prebiotic selective pressure, driven by maternal milk and infant glycan composition^28^ that influences Bifidobacteria and broader gut microbiota composition in infants.

In contrast to our finding that mother-infant FUT2+/FUT3-phenotype predicted the presence of *B. longum* subspecies *infantis* cluster strains, a prior study reported that maternal or infant FUT2 or FUT3 status did not predict the abundance of subspecies *infantis* or *longum* in children^63^. However, we included infant age as a covariate in our models, while the prior study did not^63^. In addition, to accommodate the fact that FUT2 and FUT3 work in concert biologically to produce different HBGAs and HMOs (Figure S1), we defined combined FUT2 and FUT3 phenotypes. Another smaller study found infants born to FUT2+/FUT3-mothers had lower bifidobacterial operational taxonomic unit diversity compared to other infants^39^. This study provides additional evidence that maternal FUT2+/FUT3-phenotype is associated with bifidobacterial taxon composition in the infant gut. Furthermore, we considered paired maternal and infant phenotypes^63^. More research is needed to clarify the roles of mother-infant FUT2/FUT3, maternal HMOs and the infant glycobiome as drivers of infant gut microbiome composition and development.

In our PCoA model, infant microbiome species maturation predominantly reflected decreased abundance of *B. longum* and increased abundance of *P. copri*, *F. prausnitzii*, *D. longicatena*, and *D. formicigenerans*. These species were also important predictors of infant age in a microbiome-age model previously developed from this same cohort^64^. Age-discriminatory taxa in the same genera were also identified in previously reported microbiome-age models^14^. Delayed gut microbiota-age has been reported in children with severe acute malnutrition (SAM), while improvements in microbiota-age and relative abundance of age-discriminatory taxa have been correlated with better growth^14^. Our analyses complement this literature with evidence that infant gut microbiome species maturation increased the effect of the IYCF intervention on stunting at 18mo, while greater relative abundance of *B. longum*, an age-discriminatory taxon which is associated with younger age, reduced the effect. This suggests that delayed maturation of the gut microbiome at the point when complementary foods are introduced and IYCF is initiated, may impair IYCF-induced effects on growth.

Infant age was the strongest determinant of *B. longum* relative abundance over time. This is consistent with previous reports^65^. In breastfed infants, Bifidobacteria are the most abundant gut bacteria^65,66^. Human breast milk is rich in oligosaccharides, which Bifidobacteria preferentially utilize^67^. At introduction of solid foods, a wider variety of nutrients and reduced availability of HMOs correspond to a decrease in Bifidobacterium,^68^ greater bacterial diversity and evenness, and development of a more adult-like composition by 2–3 years of age^11,69–71^.

Utilization of HMOs by Bifidobacterium varies between species and strains^72–78^. We identified three clusters of dominant *B. longum* strains that also varied by infant age and mother-infant FUT2+/FUT3-phenotype. The cluster of strains with pangenomes most closely resembling subspecies *infantis* had the highest prevalence in early infancy. However, by age 18mo there was a marked decrease in detection of *infantis* cluster strains and a corresponding increase in detection of strains with pangenomes most similar to subspecies *suis,* which had the highest prevalence at 18mo. Strains most similar to subspecies *longum* increased more slowly. This finding is consistent with a recent study that also reported a pattern of succession among three distinct gut *B. longum* clades from birth to 24mo of age. Subspecies *infantis* was dominant in early infancy but peaked at age 6mo and decreased considerably thereafter. While a transitional clade that included strains most similar to subspecies *suis* and *suillum* showed a corresponding increase in abundance that peaked by 18mo, and subspecies *longum* expanded from 15mo-24mo of age^79^. In that report, the transitional *suis*/*suillum* clade harbored functional capacity to degrade both HMOs and dietary polysaccharides, suggesting it may be an adaptation of the infant gut microbiome to a period when breastfeeding may co-occur with introduction of complementary foods^79^. We add to this work by showing that differences between mother and infant in FUT2+/FUT3-phenotype may also play a role in driving this transition, whereby, throughout follow-up, the *mother only* FUT2+/FUT3-group had a low prevalence of the *infantis* cluster and the highest prevalence of the *suis* cluster; while the *infantis* cluster was most prevalent in the *infant only* group.

*B. longum* subspecies *infantis* are particularly well adapted for HMO utilization^80,81^. *B. longum* subspecies *longum*, on the other hand, do not grow as well on HMO, and are more specialized for utilization of diet-derived polysaccharides^82^. In our analyses, UniProt gene families that were more frequent in the *infantis* cluster were more likely to be involved in HMO degradation and uptake. Two of CAZyme families more likely to be carried by the *infantis* cluster (GH29^83^ and GH33^74^) were previously reported as more prevalent in subspecies *infantis* strains. In contrast, UniProt gene families that were less frequent in the *infantis* cluster were more likely to be involved in uptake of fructose and other simple sugars, including the sugar transporter 3.A.1.2.23, which was previously described in a different *B. longum* subspecies^84^. Furthermore, strains in the *suis* and *longum* clusters were more likely to carry UniProt gene families involved in such MetaCyc pathways as pectin, xylan, and arabinogalactan degradation. UniProt gene families found more frequently in *infantis* cluster strains were also more likely to be involved in siderophore and antimicrobial biosynthesis. Heavy metals such as iron and zinc are essential minerals for nearly all bacteria and their mammalian hosts. Strategies utilized by bacteria to acquire heavy metals include siderophore biosynthesis. Siderophores are low molecular weight iron-chelating compounds that are synthesized by bacteria to scavenge iron and other essential metals such as zinc under nutrient-restricted conditions^85^. For example, Bifidobacterium species isolated from iron-deficient children efficiently sequester iron via siderophore production^86^. Siderophores may provide a competitive advantage to Bifidobacteria^87^, which along with antimicrobial biosynthesis^88^, may also help protect the infant gut from enteric pathogens that require essential metals for colonization. However, essential metals such as iron and zinc are key components in SQ-LNS formulations^89–91^, and produce improvements in linear growth and reductions in stunting risk^92,936^. While siderophore activity varies considerably between Bifidobacterium species and strains, no research to date has investigated sequestration of essential metals by subspecies *infantis* strains commonly found in resource-limited settings. Overall, our analyses suggest that greater abundance of *B. longum*, at the time when IYCF is started, characterized by a predominance of strains with carriage of gene families which confer greater capacity for HMO utilization and siderophore biosynthesis, is associated with lower impact to reduce stunting at 18mo. These suggest potential mechanisms by which a “younger” microbiome may constrain the beneficial effects of an SQ-LNS intervention on infant stunting. However, more research is required to fully elucidate the biological mechanisms and downstream pathways to stunting that could be involved.

Our results are in contrast to a previous RCT that investigated infant microbiome composition as a modifier of SQ-LNS impact on infant growth In Malawi^48^. However, in the primary analyses of that RCT, there was no effect of SQ-LNS on linear infant growth^94^. Furthermore, SQ-LNS was provided to both mothers during pregnancy and infants starting at 6mo postpartum, active control interventions (iron-folate or multiple micronutrient supplements) were provided to mothers^48,94^, and 16S rRNA gene amplification and sequencing were used to characterize the microbiota which can affect both taxon detection (including Bifidobacterium) and study results^95^. A second RCT reported evidence for effect modification of iron supplementation on growth by maternal FUT2 status, where infants of FUT2-mothers randomized to iron supplementation showed greater declines in LAZ than infants of FUT2+ mothers^96^. In addition, a third, recent RCT found that suppression of *B. longum* by amoxicillin allowed the gut microbiota of children with SAM to better adapt to a solid-food diet by reducing the abundance of taxa specialized for breast milk utilization, resulting in improved anthropometric indicators of infant nutritional status^97^. However, it is essential to note that *B. longum* subspecies *infantis* is a critical early infant gut bacteria with important benefits for infant health, including protection from enteric pathogens^98^, immune system development^99^, and reducing asthma risk^100^. *B. longum* subspecies *infantis* also improved ponderal growth when administered to infants ∼4mo of age with SAM in two RCTs^101,102^. However, the effects on LAZ were not statistically significant. Our work complements this literature. Our findings suggest that a shift away from a “less mature” microbiome, characterized by a high abundance of *B. longum* and carriage of strains that are better suited to HMO metabolism, may be critical to support efficacy of nutrient supplements that start with the introduction of complementary feeding; and we identify maternal drivers of early infant *B. longum* abundance and strain carriage which, if further elucidated, may be employed to shape the infant microbiome into a more favorable composition at this critical time in infancy.

There are some limitations of this work. First, from our cohort of 1169 HIV-unexposed infants, we had microbiome data on 172 infants. However, infants included in our analyses were comparable to infants who excluded (Table S6). Also, mother-infant FUT2+/FUT3-phenotype, which was an important determinant of *B. longum* abundance and strain cluster detection, modified the impact of IYCF on stunting in the larger sample of 792 infants without measured microbiome data in a way that was consistent with our microbiome results. That said, our small sample size may have limited power to detect effect modification by other taxa that are indicative of a “more mature” post-weaning microbiome. Our analyses were not able to investigate effect modification by actual microbial metabolic activity, maternal HMO composition or infant gut glycobiome, which would require multi-omics approaches such as metatranscriptomics and metabolomics. Finally, it is possible that the delay in microbiome maturation we characterized may reflect some reverse causality, where sick or non-thriving children are put to the breast more with higher relative breastmilk intake. However, this behavior would need to systematically occur less often in the subset of mothers and their infants who were discordant for FUT2+/FUT3-that we describe to fully explain our results, thus we think this explanation is unlikely.

In conclusion, we present analyses of moderators of IYCF impact on infant stunting at 18mo in an RCT of IYCF using SQ-LNS in rural Zimbabwe. We report that (i) infant microbiome species maturation, characterized by a shift from *B. longum* dominance, particularly *B. longum* strains that are most similar to the proficient HMO utilizer subspecies *infantis*, was associated with increased IYCF reduction of stunting risk at age 18mo; (ii) discordance in mother-infant FUT2+/FUT3-phenotype, where the mother had the phenotype but the infant did not, was associated with increased IYCF reduction of stunting risk; and (iii) reduction in *B. longum* relative abundance was also determined by mother-infant FUT2+/FUT3-phenotype. Future work should investigate how differences between maternal HMOs and infant gut glycans determine gut Bifidobacterium species and strain composition, and investigate how to tailor interventions at the introduction of complementary feeding to balance infant nutritional needs with age-related microbiome composition and function to prevent stunting.

## Methods

### Study design and participants

The Sanitation Hygiene Infant Nutrition Efficacy (SHINE) trial was a 2×2 factorial cluster-randomized trial that enrolled 5280 pregnant women at a median age of 12.5 weeks gestation between November 2012 and March 2015 to test the impact of improved household water quality, sanitation, and hygiene (WASH) and improved infant and young child feeding (IYCF) via provision of SQ-LNS to the infant from age 6mo-18mo, on linear growth and anemia at age 18mo. A detailed description of the SHINE trial design and methods has been published^51,103^.

Briefly, research nurses made home visits twice during pregnancy and at infant ages 1, 3, 6, 12, and 18 months. At baseline, maternal education and age, household wealth^104^, existing water and sanitation services, and household food security^105^ were assessed, and mothers were tested for HIV via a rapid testing algorithm. Infant birth date, weight, and delivery details were transcribed from health facility records. Gestational age at delivery was calculated from the date of the mother’s last menstrual period ascertained at baseline. Infant weight, length, and mid-upper arm circumference were measured at every postnatal visit. Nurses were standardized against a gold-standard anthropometrist every 6 months, with retraining provided to those who failed to meet predefined criteria.

Part-way through the trial (from mid-2014 onwards) mother-infant pairs were invited to join a substudy to collect additional biological specimens. Women were informed about the substudy at their 32-week gestation visit and those with live births were enrolled at the 1-month postnatal visit, or as soon as possible thereafter^106,107^.

### Specimen collection and processing

Mothers collected fecal specimens prior to the research nurse visit. Fecal specimens were placed in a cold box and transported to the field laboratory, where they were stored at –80°C until transfer to the central laboratory in Harare for long-term archiving at –80°C, with generator back-up. Fecal specimens were transferred via private courier on dry ice from Harare, Zimbabwe to Vancouver, British Columbia for metagenomic analyses.

The Qiagen DNeasy PowerSoil Kit was used to extract total DNA from 200mg of feces, according to manufacturer’s instructions. Paired-end libraries were constructed using the Illumina TruSeq kit and using New England Biosystem TruSeq compatible library preparation reagents. Libraries were sequenced at the British Columbia Genome Sciences Centre using the Illumina HiSeq 2500 platform. Forty-eight libraries were pooled and included per sequencing lane. Negative controls were included to capture microbial contamination in the DNA extraction and library preparation steps.

The analyses presented here utilize data and specimens from HIV-uninfected mothers and their infants enrolled in the specimen collection substudy. The fecal microbiome was characterized in 354 specimens collected from 172 HIV-unexposed infants from 1 to 18mo of age. A mean(sd) of 2.0(1.0) samples were analyzed per child. Infants included in these analyses largely resembled the population of live-born infants from the wider SHINE trial who were not included in these analyses (Table S6). However, infants in the current analyses had slightly older mothers and longer gestational ages, and fewer were born during the hungry season, but more were in a household that met the minimum dietary diversity score (Table S6). Overall, the majority were born by vaginal delivery (91.6%) in an institution (91.5%) and were exclusively breastfed (83.2% at 3 months). The prevalence of stunting was 26.9% at 18mo in this sub-study (Table S6).

### Bioinformatics

Sequenced reads were trimmed of adapters and filtered to remove low-quality, short (<60 base-pairs), and duplicate reads, as well as those of human, other animal or plant origin using KneadData with default settings. Overall, 354 unique whole metagenome sequencing datasets were used from fecal specimens collected from 1mo to 18mo from 172 infants with available mother and infant FUT2 and FUT3 phenotypes (Figure S2). On average, 10.8±3.7 million paired end quality-filtered reads were generated per sample. Assessment of negative controls and technical variation have been previously reported^64^. Species composition was determined by mapping reads to clade-specific markers using MetaPhlAn3, while functional gene and metabolic pathway composition was determined using HUMAnN3 against the UniRef90 database, both with default settings^52^. Bacterial species and pathway abundance estimates were normalized to relative abundances. UniProt gene family profiles were generated for dominant *B. longum* strains in fecal metagenomes with sufficient coverage for pangenome analysis using PanPhlan3^52^. To facilitate interpretation of UniProt gene family profiles, the minimum set of biological pathways sufficient to explain the gene families identified in each strain was determined using MinPath with default settings^58^ and the MetaCyc database^57^.

### Assessment of FUT2 and FUT3 status

Saliva samples were collected by oral swab from mothers and infants. Available saliva from any follow-up visit was selected to assess FUT2 and FUT3 status. Secretor versus non-secretor (FUT2) and Lewis-positive versus Lewis-null (FUT3) status were ascertained for infants and their mothers using a previously reported phenotyping assay^108^. We defined FUT2 and FUT3 phenotype combinations as Lewis-positive non-secretors (FUT2-/FUT3+), Lewis-positive secretors (FUT+/FUT3+), or Lewis-null secretors (FUT2+/FUT3-) (Table S11) according to the histo-blood group antigen synthesis pathways defined in Figure S1. Paired mother-infant phenotype concordance or discordance was defined as presented in Table S1.

Of 1,169 mother-infant pairs, 999 (85.4%) mothers and 1104 (94.4%) infants were tested. Of these, FUT2 or FUT3 status could be determined in 889 (76.0%) and 999 (85.4%) mothers and infants, respectively (Table S11). Of those whose FUT2 or FUT3 status could be ascertained, secretors were the most frequent FUT2 phenotype (88.1% and 85.6% in mothers and infants respectively), while Lewis-positive was the most frequent FUT3 phenotype (76.8% and 74.9% in mothers and infants, respectively) (Table S11). Most mothers and infants were Lewis-positive secretors (60.5% and 64.9%, respectively), followed by FUT2+/FUT3-(25.1% and 23.2%, respectively), and Lewis-positive non-secretors (14.4% and 11.9%, respectively) (Table S1).

### Statistical analyses

Infant characteristics that explain species β-diversity were evaluated using constrained principal coordinates analysis (PCoA) of Bray-Curtis dissimilarities (*capscale*)^109^. Characteristics of interest included factors that are known to be correlated with microbiome composition (infant age, sex, EBF at 3mo, dietary diversity, maternal and infant FUT2 and FUT3 phenotype). Statistical significance was tested by permutational analysis of variance using distance matrices with 1000 permutations (*adonis2*)^110^. We then developed a final multivariable constrained PCoA model that included covariates which explained a significant fraction (p<0.05) of the variance in microbiome composition (infant age at specimen collection), as well as mother-infant FUT2+/FUT3-discordance given our *a prior* hypothesis that FUT2 and FUT3 phenotypes are important determinants of infant microbiome composition and our finding that mother-infant FUT2+/FUT3-discordance was an important modifier of IYCF efficacy to reduce stunting at 18mo. Constrained PCoA axis scores in the final multivariable model represented changes in microbiome composition (species turnover) along gradients defined by each infant characteristic included in the full model.

Since the IYCF intervention was started at the 6mo follow-up visit, we assessed interaction between randomization to IYCF and mother-infant FUT2+/FUT3-discordance or microbiome composition using data from the 6mo visit as covariates. The primary outcome was stunting at 18mo and LAZ at 18mo was a secondary outcome. We fitted separate models for stunting and LAZ at 18mo. Interaction was assessed on the additive risk difference scale, which is most appropriate for statistical estimation of synergistic biological effects^111^. We used generalized linear models (*glm*) with a Gaussian distribution, an identity link, sandwich standard errors (*sandwich*)^112^ and an IYCF-by-mother-infant FUT2+/FUT3-phenotype interaction term. We repeated these analyses using IYCF-by-PCoA axis score interaction terms, where PCoA axis scores were derived from the final multivariable constrained PCoA model. One model was fitted per constrained PCoA axis. For species that were strongly associated with PCoA axis scores (loadings > 0.5 or < –0.5), analyses were repeated to assess IYCF-by-species interactions. We fitted a separate model for each species of interest. Models also included IYCF, infant sex, mother-infant FUT2+/FUT3-discordance, infant age at specimen collection, an indicator of whether infants met the minimum dietary diversity score at specimen collection, and LAZ at specimen collection. We did not include WASH arm because, in prior analyses, the SHINE WASH intervention did not affect stunting or LAZ at 18mo^51^ nor infant gut microbiome composition^64^. P-values were adjusted for multiple hypothesis testing to preserve the false discovery rate^113^.

### Identification and analysis of Bifidobacterium longum strain clusters

*B. longum* strain profiles produced with PanPhlan3.0, which indicate whether UniProt gene families are present in a strain^114^, were converted to Jaccard dissimilarity matrices and visualized by PCoA to ascertain the existence of strain clusters (*capscale*). Three clusters were identified, and strain cluster membership was determined by hierarchical clustering of Jaccard dissimilarities and Ward’s error sum of squares algorithm (*hclust*)^115^. Hierarchical clustering dendrograms were cut at a height to obtain three clusters.

Differences in UniProt gene family profiles between *B. longum* strain clusters were determined by two-sided Fisher’s Exact test (*fisher.test*), with adjustment for multiple hypothesis testing to preserve the false discovery rate^113^, and were visualized using heatmaps (*heatmap3*). 3260 UniProt gene families were differentially present between strain clusters after FDR-adjustment. We performed overrepresentation analyses^53^ using one-sided Fisher’s Exact tests (*fisher.test*) to determine whether the differentially present gene families were more likely to function as particular CAZymes^55^ or transporters^56^, or in specific GO biological processes^54^. These analyses were repeated using the biological pathways determined with MinPath^58^. 115 pathways were differentially present between strain clusters after FDR-adjustment. We performed overrepresentation analyses of these pathways to determine whether they were more likely to have particular biological functions defined by MetaCyc pathway types.

### Predictors of Bifidobacterium relative abundance and strain detection

Predictors of *B. longum* relative abundance over time were assessed using mixed-effects zero-inflated beta regression estimated by restricted maximum likelihood (*gamlss*)^116^. The model included infant age at specimen collection, sex, EBF at 3mo, minimum infant dietary diversity at specimen collection, and mother-infant FUT2+/FUT3-discordance, with random intercepts (*re*) and a first order autocorrelation structure (*corCAR1*).

Predictors of *B. longum* strain cluster were assessed by logistic regression (*glm*) using an indicator of strain cluster presence as the dependent variable, with the same covariates, and sandwich standard errors. An individual model was fitted separately for each cluster. Bias corrected^117^ logistic regression was used to facilitate stable parameter estimation due to separation resulting from the small sample size^118^.

All statistical analyses were conducted in R version 4.2.0. PCoA and adonis2 were performed using the *vegan* package^119^. Heatmaps were generated with *heatmap3*. Mixed-effects zero-inflated beta regression was performed using the *gamlss* package^120^. Bias corrected logistic regression models were fitted using the *brglm2* package. Sandwich standard errors were generated with the *sandwich* package.

## Resource availability

### Lead contact

Further information and requests for resources and reagents should be directed to and will be fulfilled by the lead contact, Ethan Gough.

## Materials availability

This study did not generate new unique reagents.

## Data and code availability

The raw metagenome sequencing data generated in this study have been deposited in the European Bioinformatics Database under accession code PRJEB51728. Final processed and annotated metagenome sequencing data files (taxa and pathways) are available at https://doi.org/10.5281/zenodo.7471082. Epidemiologic data files are available at Code for statistical analyses are available from the corresponding author upon request.

## Supplemental Tables

**Table S8. Overrepresentation of UniProt gene families and Metacyc pathways, which differ between strain clusters, by GO biological process, CAZyme or Transporter class or Metacyc pathway type.**

## Ethics approvals

All SHINE mothers provided written informed consent. The Medical Research Council of Zimbabwe (MRCZ/A/1675), Johns Hopkins Bloomberg School of Public Health (JHU IRB # 4205.), and the University of British Columbia Ethics Board (H15-03074) approved the study protocol, including the microbiome analyses. The SHINE trial is registered at ClinicalTrials.gov (NCT01824940).

## Data Availability

All data produced in the present study are available upon reasonable request to the authors.

## Acknowledgments

We thank all the mothers, babies, and their families who participated in the SHINE trial and all members of the SHINE trial team (all members listed here: https://doi.org/10.1093/cid/civ844). We particularly thank the leadership and staff of the Ministry of Health and Child Care in Chirumanzu and Shurugwi districts and Midlands Province (especially environmental health, nursing, and nutrition) for their roles in operationalization of the study procedures, the Ministry of Local Government officials in each district who supported and facilitated field operations, Phillipa Rambanepasi and her team for proficient management of all the finances, Virginia Sauramba for management of compliance issues, and the programme officers at the Bill & Melinda Gates Foundation and the Department for International Development (UK Aid), who enthusiastically worked with us over a long period to make SHINE happen.

Funding was from the Bill & Melinda Gates Foundation (OPP1021542 and OPP1143707; J.H.H. and A.J.P.), with a subcontract to the University of British Columbia (20R25498; A.R.M.). United Kingdom Department for International Development (DFID/UKAID; J.H.H. and A.J.P.). Wellcome Trust (093768/Z/10/Z, 108065/Z/15/Z, 206455/Z/17/Z, 203905/Z/16/Z and 210807/Z/18/Z; A.J.P., R.C.R. and C.E.). Swiss Agency for Development and Cooperation (J.H.H. and A.J.P.). US National Institutes of Health (2R01HD060338-06; J.H.H.). UNICEF (PCA-2017-0002; J.H.H. and A.J.P.). The Nutricia Research Foundation (2021-52; E.K.G.) The funders had no role in the design of the study and collection, analysis, and interpretation of data and in writing the manuscript.

## Author contributions

A.R.M., L.E.S., R.J.S., M.N.N.M., J.H.H. and A.J.P. conceptualized and designed the study. K.M., R.N., B.C., F.D.M., N.V.T., J.T., and B.M. collected data and biospecimens. H.M.G., I.B., S.K.G., R.C.R., F.F. and L.C. processed fecal specimens. M.K. conducted laboratory analyses to ascertain FUT2 and FUT3 status. C.E., J.C. and E.K.G. developed the FUT2 and FUT3 analysis plan. E.K.G. developed and conducted the microbiome statistical analysis plan. T.J.E. and E.K.G. conducted bioinformatics. A.R.M. and E.K.G. analyzed and interpreted the data. E.K.G. wrote the original manuscript draft. All authors reviewed the manuscript. A.R.M., A.J.P. and J.H.H. supervised and verified the data.

---

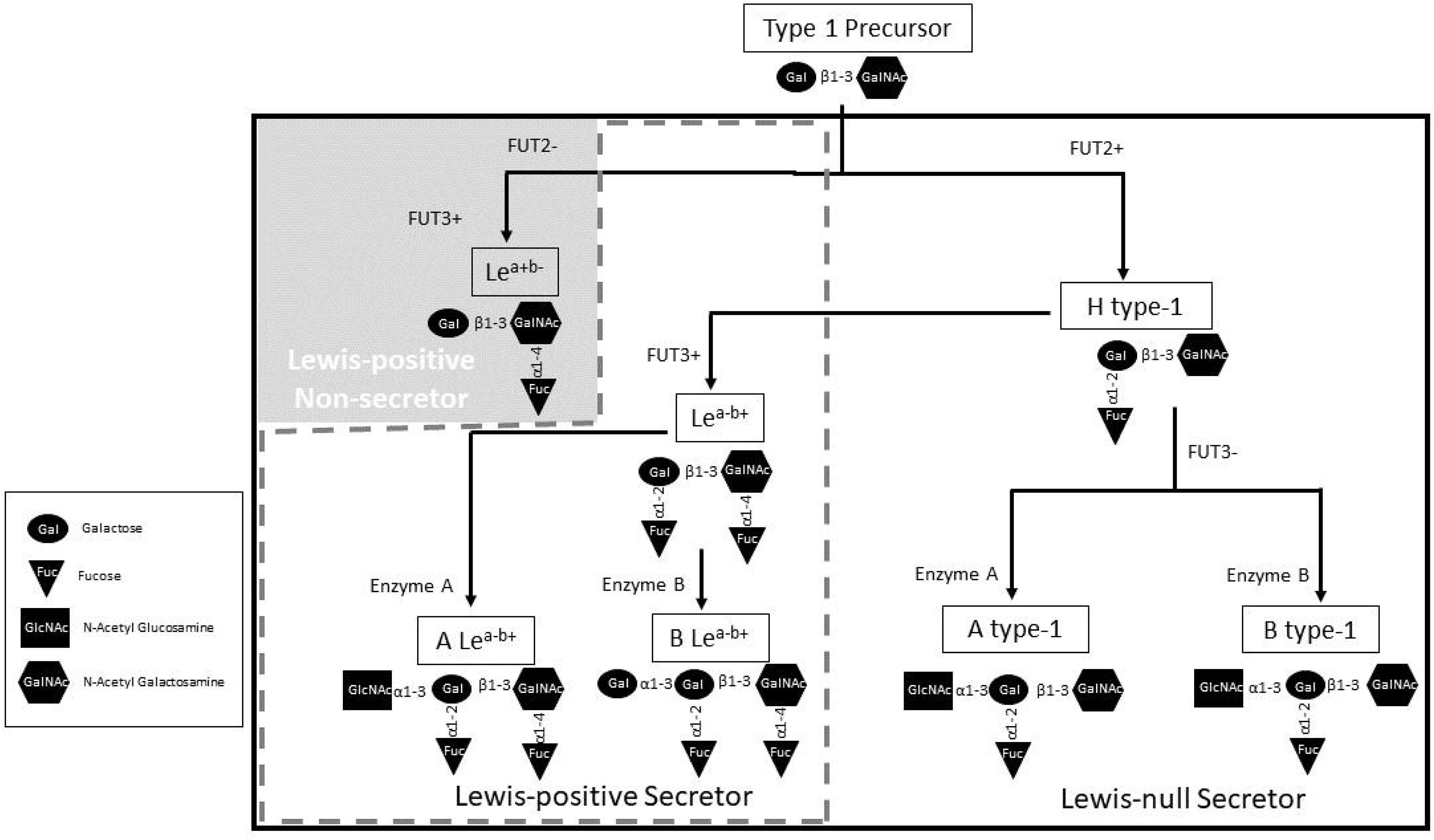

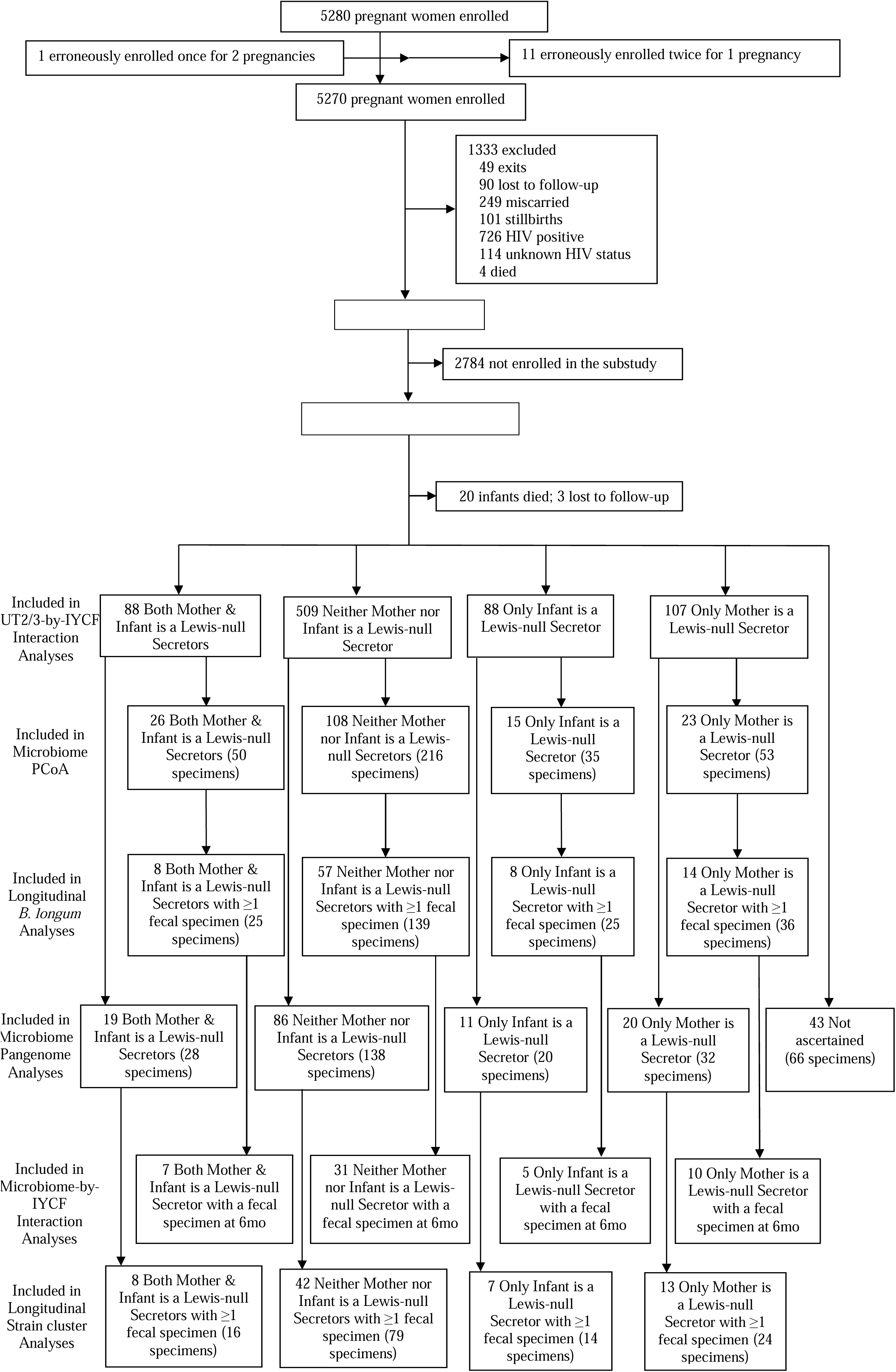

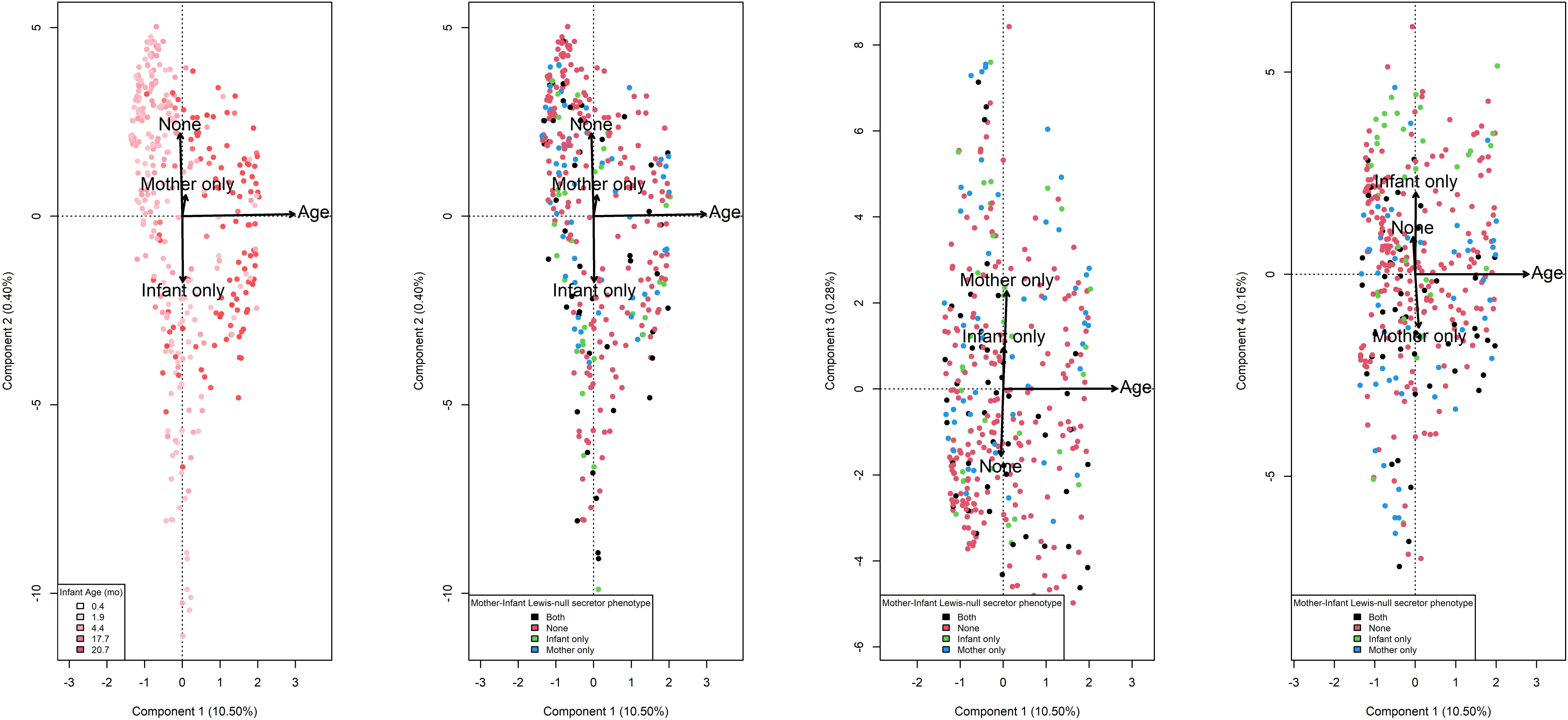

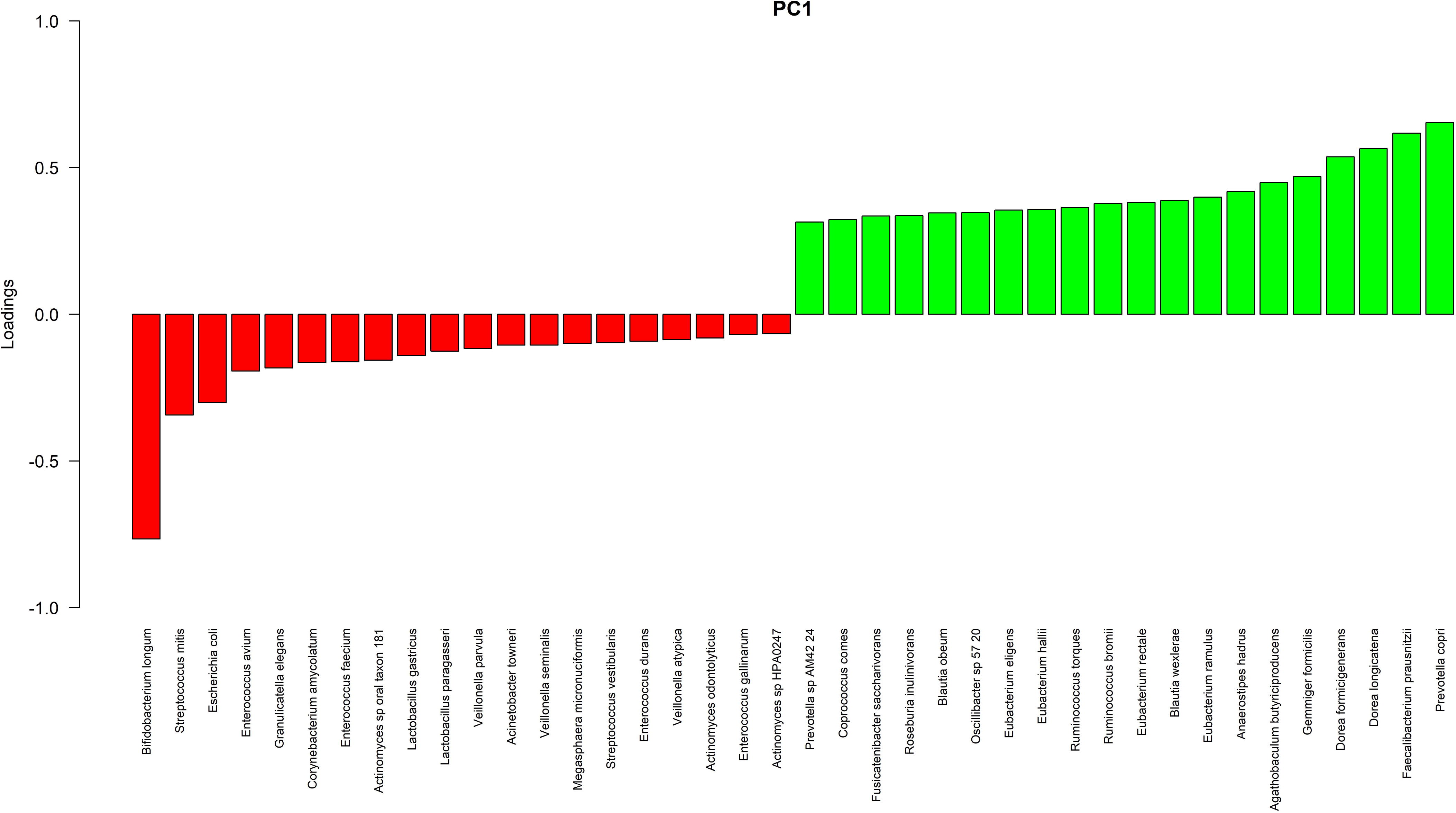

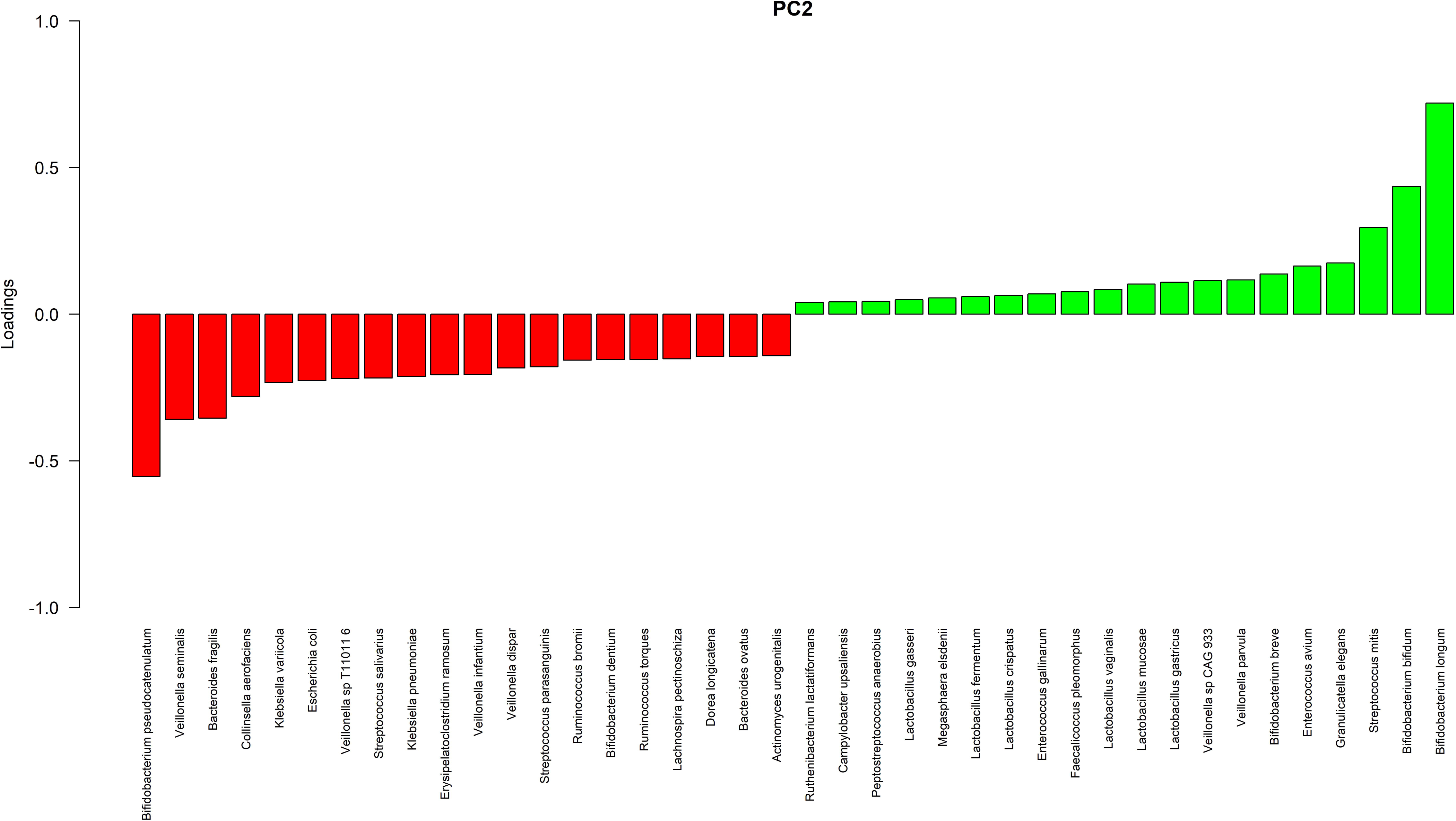

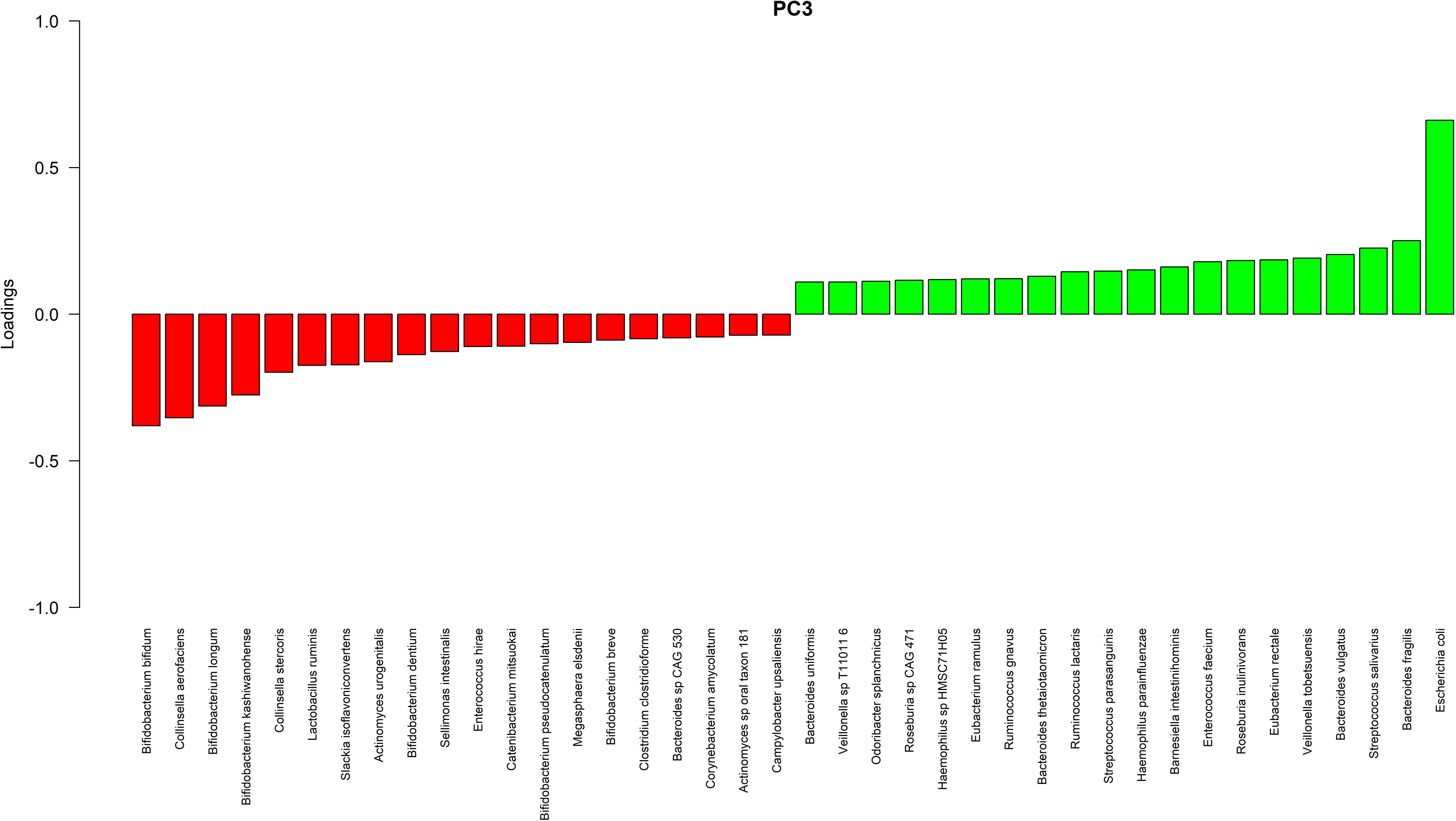

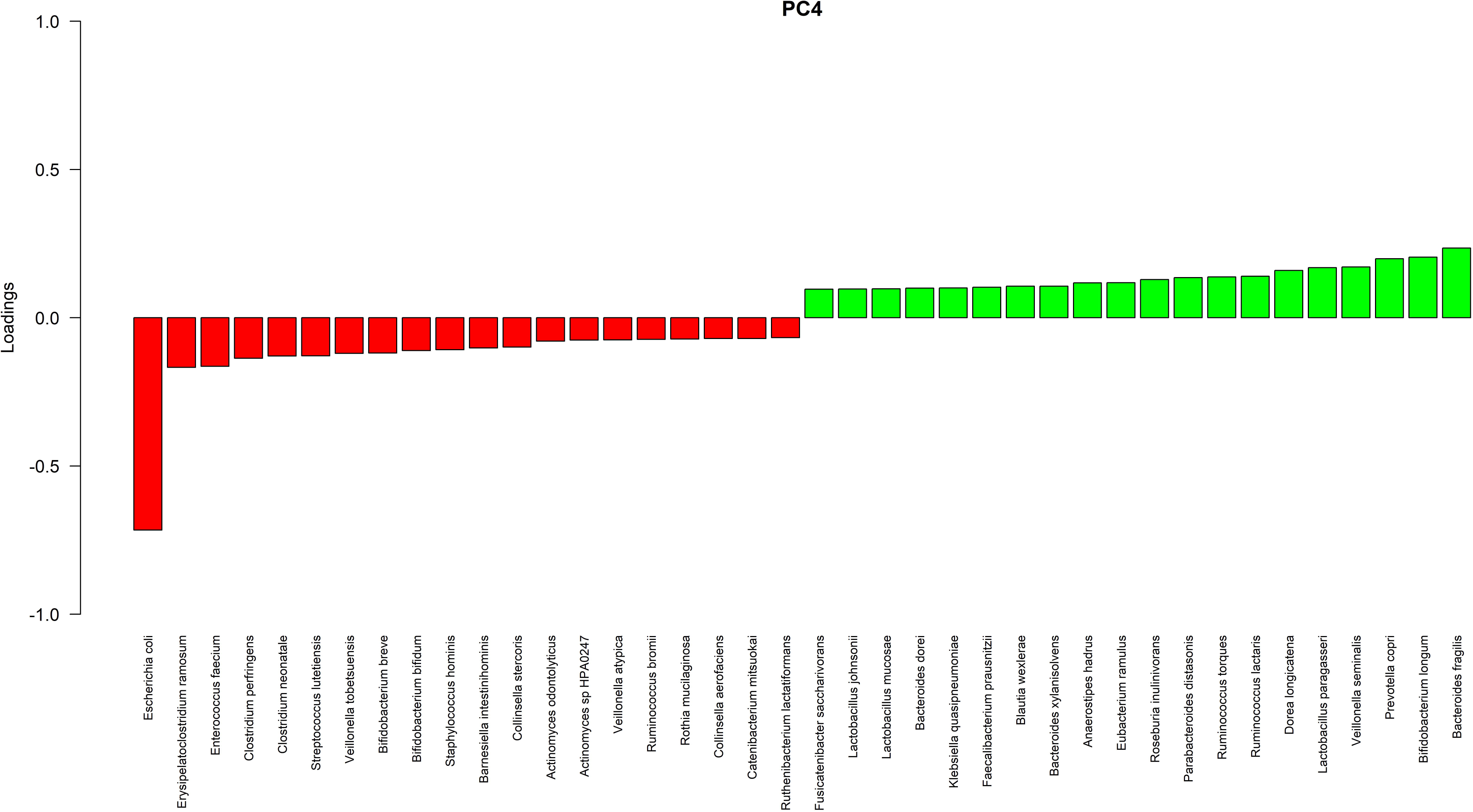

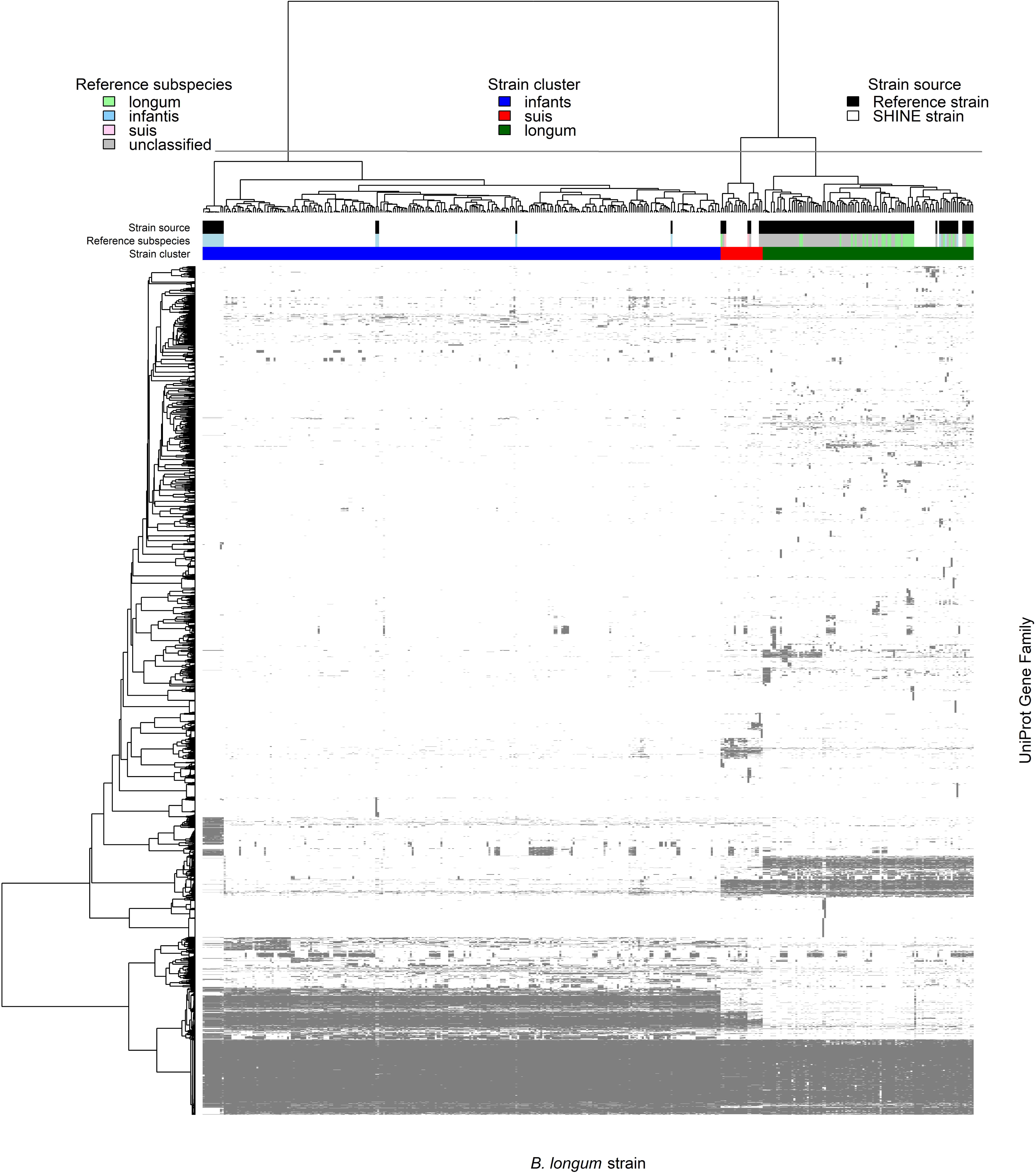

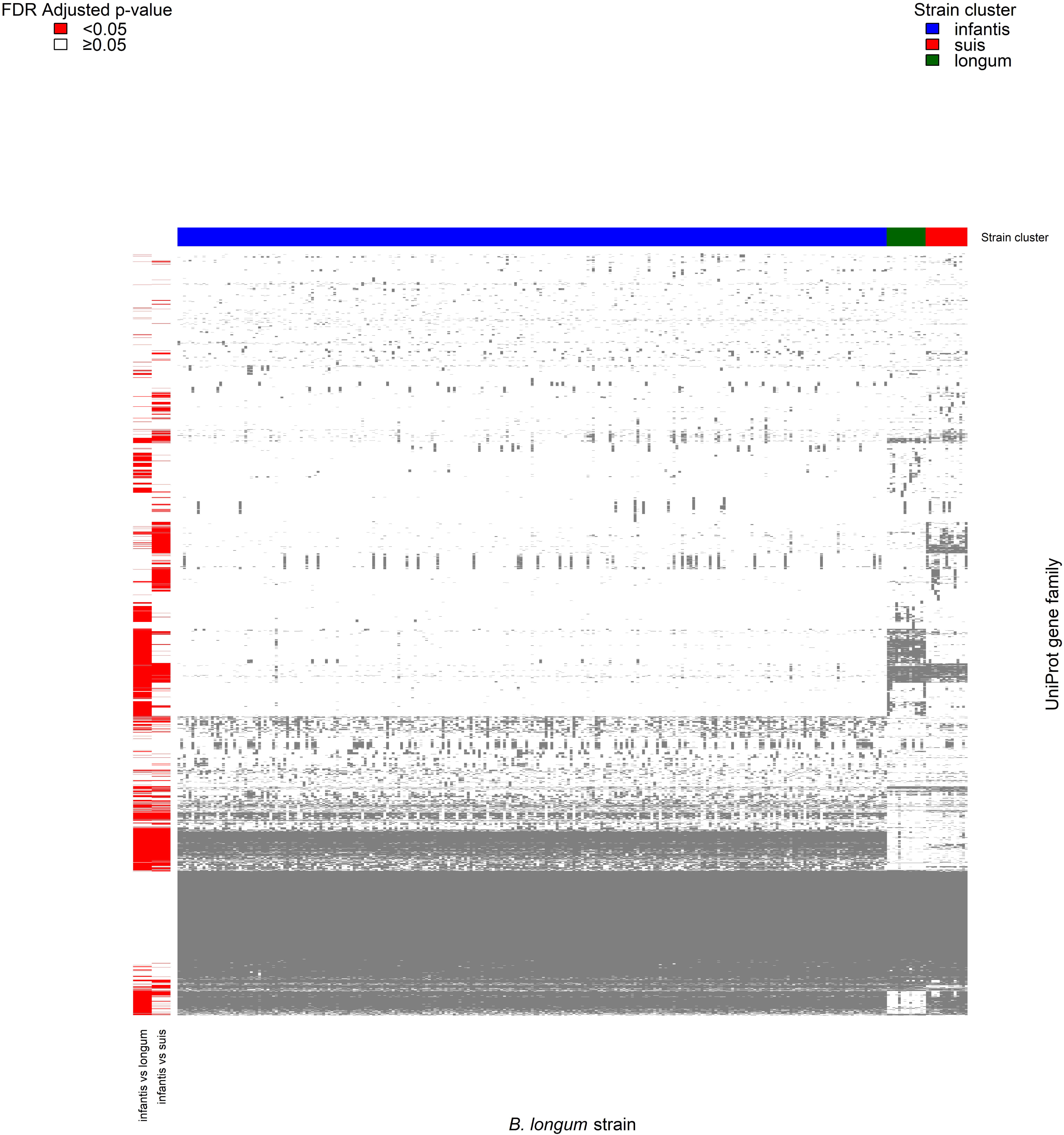

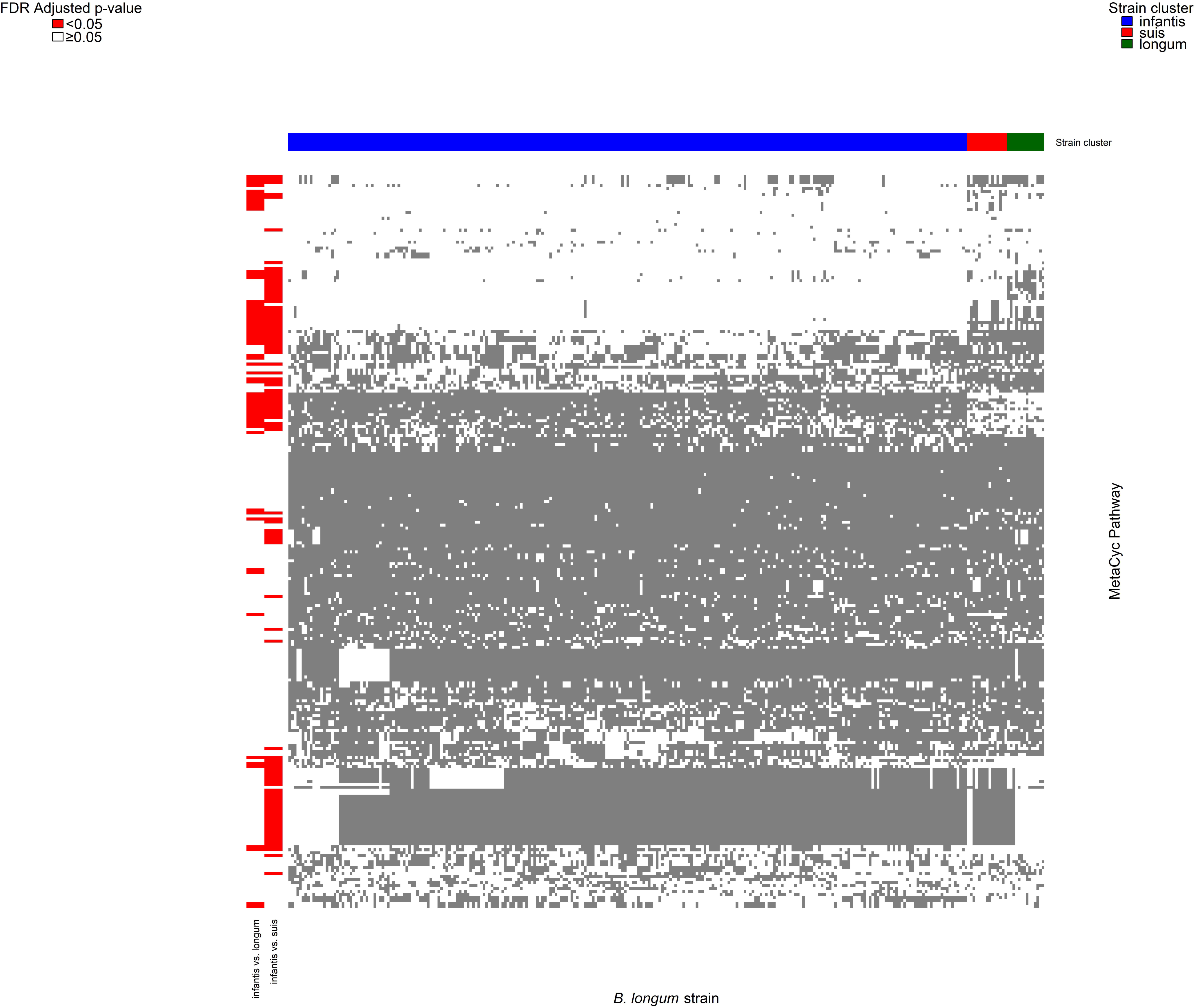

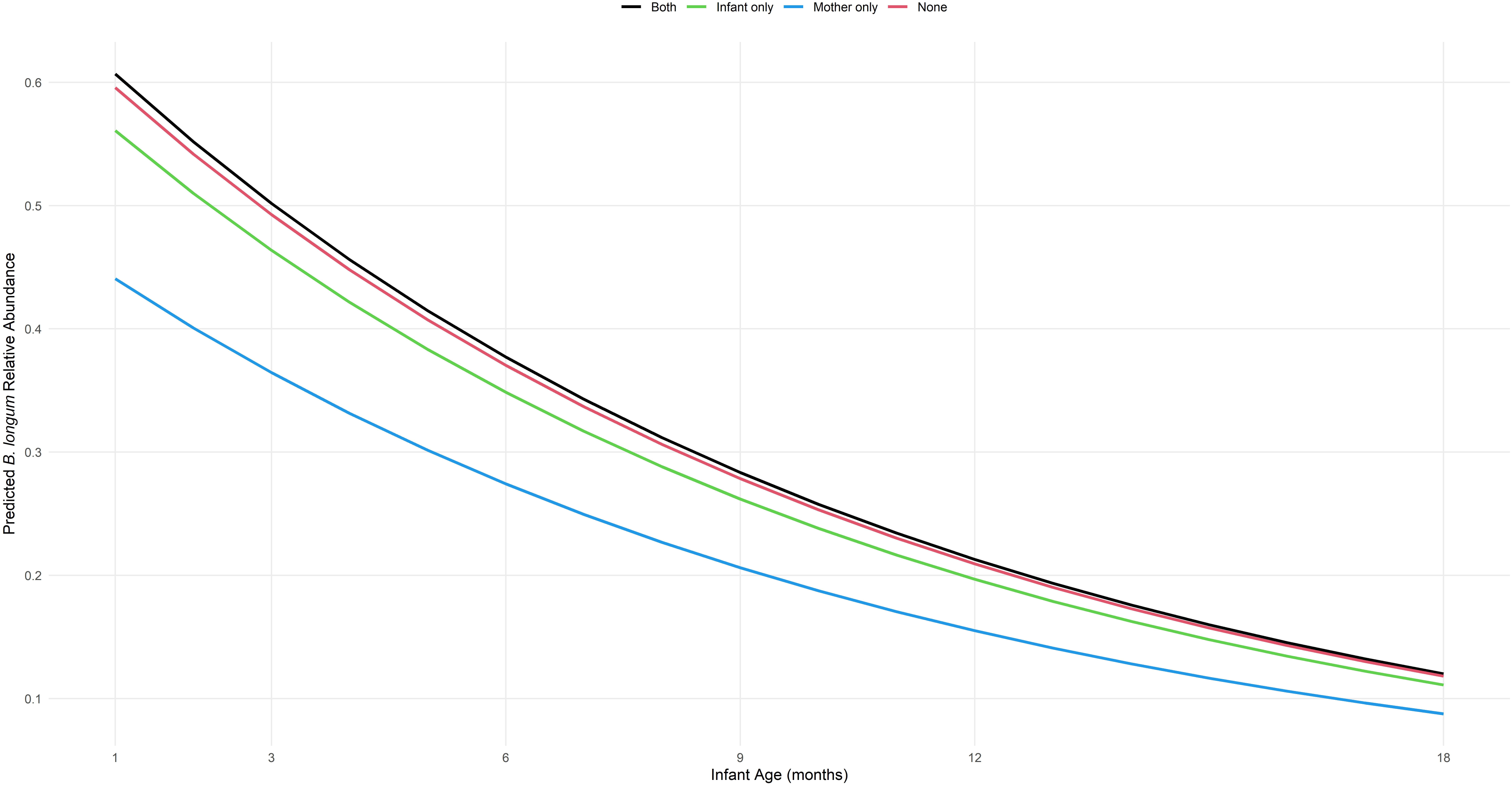

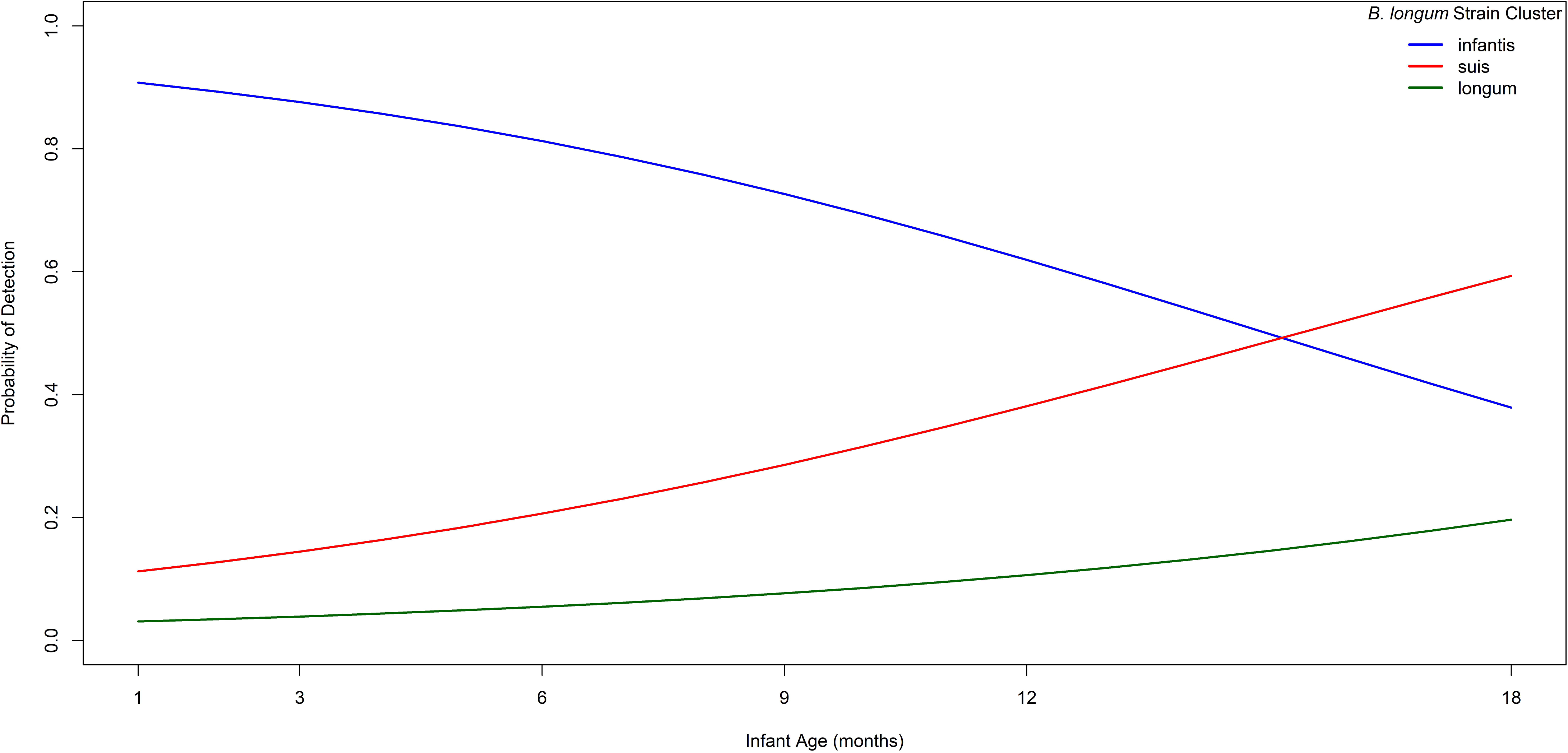

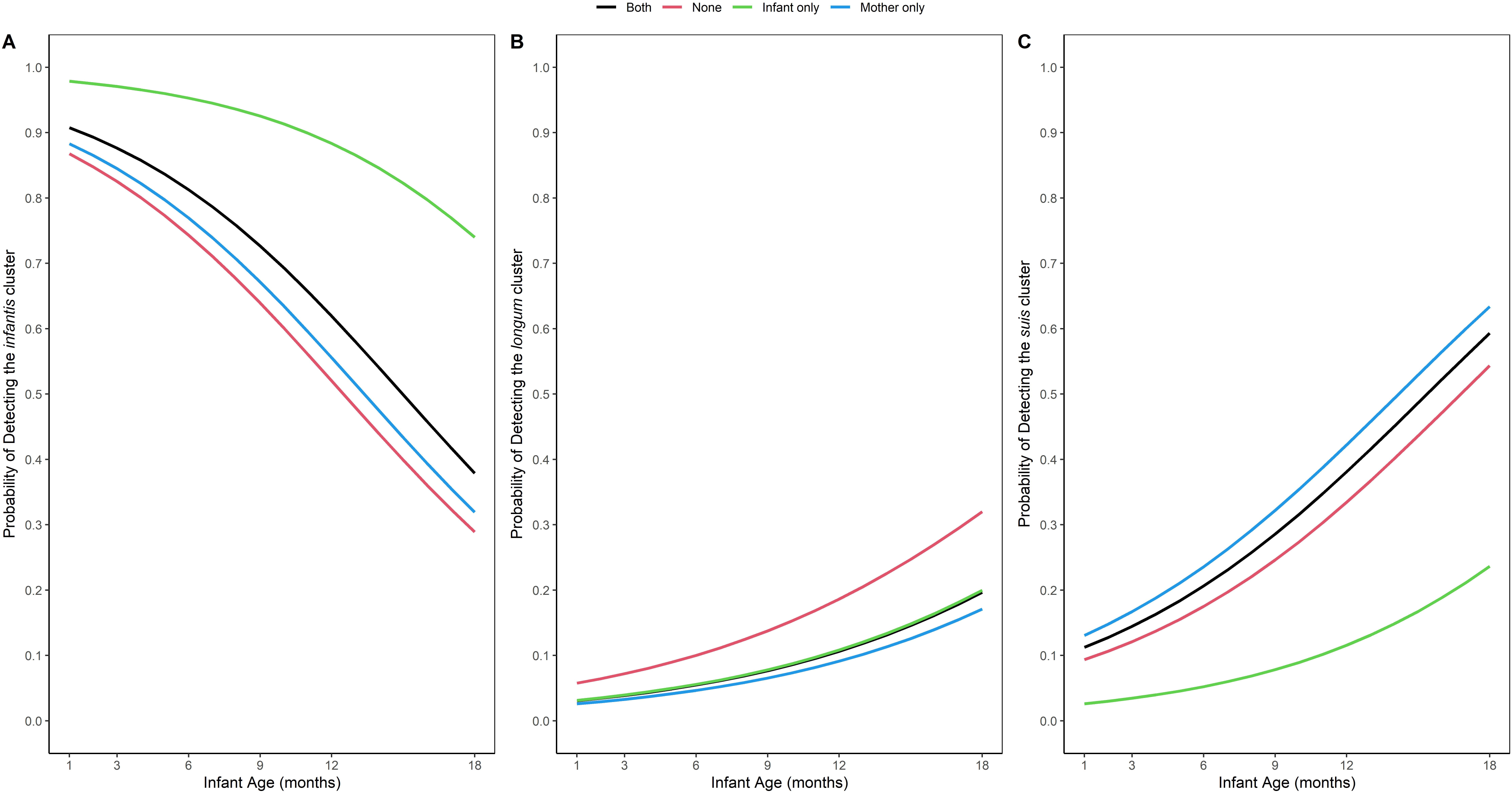

